# Post Stroke Cognitive Impairment: more than a lesion-symptom model

**DOI:** 10.1101/2025.02.19.25322521

**Authors:** Hanne Huygelier, Margaret Jane Moore, Annick Odom, Nora Tuts, Hella Thielen, Mauro Mancuso, Céline R. Gillebert, Nele Demeyere

## Abstract

Post-stroke cognitive impairment (PSCI) is highly heterogeneous, reflecting both domain-specific deficits associated with focal lesions and broader impairments linked to premorbid health and demographic factors. In the current study we investigated whether PSCI can be distinguished into distinct across-domain cognitive profiles and how such PSCI profiles are associated with lesion neuroanatomy, demographic and premorbid health factors.

This cross-sectional study employed data-driven analyses (i.e., Latent Class Analysis) on 2172 stroke survivors who completed a domain-specific cognitive screen (i.e., Oxford Cognitive Screen) within 6 months after stroke (M = 19; Mdn = 7, SD = 35.7 days). In addition, the association of the PSCI profiles with lesion and demographic characteristics was investigated.

We identified two viable cognitive class solutions: a 5-class model capturing classical and novel PSCI profiles and a more detailed 13-class model reflecting a broader set of distinct cognitive profiles that commonly occur after stroke. The 5-class solution distinguished classical lateralized deficits (e.g., aphasia, neglect) alongside a minimal impairment and non-lateralized global impairment profile. In contrast, the 13-class solution provided finer-grained differentiation, particularly for non-lateralized cognitive profiles which were more strongly associated with premorbid health and education level. Importantly, lesion anatomy alone could not fully account for class distinctions. While lesion location was predictive, particularly, in hyper-acute stages, profiles for patients tested 2 weeks post-stroke revealed less influence of lesion location and more of lesion volume.

These findings underscore the importance of considering both anatomical and premorbid factors in understanding PSCI. By providing a nuanced classification of PSCI profiles, this study establishes a foundation for future translational research aimed at improving clinical care and predicting cognitive trajectories.

## Introduction

Cognitive impairment is a common consequence of stroke which is associated with poor recovery and outcomes in daily life.^1^ While theoretical neuropsychology has traditionally emphasised selective deficits associated with damage to specific brain regions, the clinical reality is that post-stroke cognitive impairments (PSCI) rarely occur in isolation. Indeed, PSCI typically involves co-occurring impairments across multiple cognitive domains.^2–5^ Past research has demonstrated that patterns of cognitive associations (and dissociations) vary widely across individuals.^6–8^ While capturing this complexity is crucial for understanding the cognitive needs of patients, it is equally important to explore whether individual variability in PSCI can be attributed to a smaller set of underlying factors.

To this end Corbetta et al. (2015) evaluated language, motor, memory, and attention in first-time stroke survivors (n=67) to identify factors explaining behavioral correlations across subjects. They found 3 factors (one lateralized to each hemisphere + a non-lateralized factor) accounting for 69% of behavioral variance.^9^ Similarly, Bisogno et al. (2021) found that approximately 50% of variance on the Oxford Cognitive Screen (OCS), a domain-specific cognitive screen for stroke patients, could be accounted for by 3 factors. The first factor captured language, calculation, praxis, right-lateralized spatial neglect, and memory. The second factor loaded on left motor and visuospatial deficits, and the third factor loaded on right motor impairment.^10^ However, a more recent larger-scale (n=1973) Principal Component Analysis (PCA) found that OCS performance was best captured by a 6-factor solution (language/arithmetic, memory, visuomotor ability, orientation, spatial exploration, and executive functions).^11^ Overall, past research has suggested that the variability in PSCI may be captured by a reduced number of dimensions, but the number and underlying nature of these dimensions have not been reliably established.

While PCA has been useful in describing patterns of PSCI associations, this approach may underestimate the complexity of PSCI.^12^ Sperber et al. (2022) demonstrated that the systematic spatial variability of stroke lesion anatomy alone is sufficient to result in an apparent low-dimensional structure underlying PSCI, even when all simulated impairments were independent. Consequently, Sperber et al. (2022) called for future studies to develop and compare latent structures of varying complexity, suggesting that additional factors which do not increase explained variance but produce an intuitively interpretable solution should be retained in solutions. It is also important to consider that profiles of lesion-induced disconnection, in addition to lesion location, may help explain patterns of PSCI associations. Past work has demonstrated that individual PSCI domain impairments can be mapped onto distinct patterns of network-level disconnection^13,14^, but the extent to which these disconnection patterns may help account for common PSCI profiles remains unclear.

However, it is important to recognize that lesion anatomy alone is unlikely to fully account for the variability in PSCI. This is because the relationship between impairments and lesion anatomy is often confounded by pre-existing neurovascular changes.^15^ Premorbid factors including cerebral atrophy^8,16^, white matter integrity^17,18^, and education level^6,19^ are each associated with an increased risk of PSCI. There is also high comorbidity between stroke and major neurocognitive disorders and mild cognitive decline^20^. These brain health markers are typically associated with impairment in memory, executive functions, processing speed and language^18,21,22^, which can occur alongside stroke-related cognitive impairments. The latter complicates the clinical picture of PSCI.

Although many studies have documented the importance of premorbid factors for PSCI, there is a lack of studies investigating across-domain PSCI profiles and how such profiles link to lesion anatomy and premorbid brain health. That is, prior studies using data-driven strategies to investigate the structure of PSCI, have either focused on separate cognitive domains^23–25^, a global cognition outcome^26–29^ or have focused on a select group of stroke patients (such as first-ever stroke patients with good premorbid brain health).^9,26–28^ Given the high comorbidities in the clinical reality of stroke, it is important to disentangle distinct cognitive profiles and investigate to what extent such profiles align with specific lesion topographies versus pre-existing neurovascular changes, especially in a clinical stroke sample.

The present study seeks to address this gap by using latent class analysis to investigate cognitive profiles in a large stroke cohort assessed with the Oxford Cognitive Screen (OCS). This approach allows to explore the structure of PSCI in a clinically representative stroke sample beyond the limitations of global cognitive screens (Demeyere et al., 2016a). Specifically, our objectives were to (1) distinguish cognitive profiles after stroke, (2) assess and compare the interpretability of these profiles, and (3) evaluate the extent to which stroke lesion anatomy, premorbid brain health and demographic variables influence the structure of PSCI.

## Materials and methods

Data of 2172 stroke patients were aggregated from Belgian, Italian and UK databases.^3,30–33^ These studies included all patients who were able to concentrate for 15-20 minutes (Supplementary Materials 1). All patients provided informed consent and all procedures followed the Helsinki declaration.

### Cognitive Screen

Domain-specific impairments were assessed with three language versions of the Oxford Cognitive Screen (i.e., OCS, OCS-IT, OCS-NL).^3,30,32–35^ These OCS translations have each been validated in stroke patients. ^3,32,33^ The OCS is designed to be inclusive for patients with common stroke-related impairments including aphasia, spatial neglect, primary visual and motor impairments.^3,30^ The OCS consists of ten subtasks indexing: language (picture naming, semantics, sentence reading), memory (orientation, verbal and episodic memory), numerical cognition (writing numbers, calculation), praxis (imitating meaningless gestures), and executive function (trail making) / attention (cancellation test). Detailed descriptions of OCS subtests are reported elsewhere.^3,30^ All impairment classifications were made based on clinical thresholds which were age-corrected for the OCS-NL^34^ and OCS-IT^35^.

### Neuroimaging data

All patients with lesion masks and behavioural data collected within 45 days of stroke onset were included in neuroimaging analyses (n = 515). Lesion maps were derived from acute (<31 days post-stroke) clinical neuroimaging (442 CT, 4 T1, 62 T2, 6 FLAIR) which were collected in the UK OCS screening programmes.^3,30^ Previous studies have found that CT and MR yield comparable lesion mapping results.^36–38^ Patients with clear evidence of multiple, temporally distinct lesions were excluded.

Lesions were manually delineated by trained experts on axial slices using MRIcron. Native-space lesion masks were smoothed at 5 mm full-width at half-maximum in the z-direction, binarized (0.5 threshold), reoriented, warped into 1×1×1mm stereotaxic space using Statistical Parametric Mapping^39^ and Clinical Toolbox^40^ functions, and were visually inspected for quality. This normalisation procedure employs age-specific CT or MR templates.^40^ Exploratory analyses were conducted to ensure behavioural results were consistent between the total sample and subset of patients with lesion data (Supplementary Materials 2).

To quantify network-level disconnectivity, lesion masks were used to estimate parcel-wise disconnectivity across all cortical areas defined in the Schaefer-Yeo Atlas (100 parcels)^41^ and subcortical/cerebellar areas in the AAL^42^ and Harvard-Oxford atlases (35 parcels). The atlases define 7 networks: Control, Default, Dorsal Attention, Limbic, Somatic Motor, Ventral Attention, and Visual networks as well as networks connecting subcortical/cerebellar structures.^43^ In cases where lesions intersect with a streamline (i.e., a connection between two parcels), the relevant streamline is considered to be disconnected.^44^ Network-level disconnectivity was calculated as the proportion of streamlines which terminate (end or begin) in each pair of parcels that were disconnected.^44^ Tract-level structural disconnection was quantified by calculating the percent of streamlines within each of the HCP-842’s 70 white matter tracts which were disconnected by lesions. These methods are standard when in-vivo tractography data is not available.^44^

Atrophy was assessed using the Global Cortical Atrophy (GCA) scale.^45^ The atrophy in 13 brain regions was assigned a score of 0 (none), 1 (mild), 2 (moderate), or 3 (severe). Where regions were obscured by stroke lesions, regions were assigned the score of the homologous region within the undamaged hemisphere. The severity of white matter lesions was determined using the Fazekas scale.^46^ Deep white matter lesions were rated as 0 (absent), 1 (punctuate foci), 2 (beginning confluence of foci), or 3 (large confluent areas). Periventricular white matter lesions were rated as 0 (absent), 1 (symmetrical caps), 2 (smooth halo), or 3 (irregular hypoattenuations extending into deep white matter). Total GCA and Fazekas scores (n = 475) were calculated by adding all region scores (GCA range = 0 – 39, Fazekas range = 0 - 6). Both scales have been validated for use in CT^47^ and MR stroke imaging^45,46^.

### Statistical analyses

Data-analysis was performed using R 4.0.2, python 3.9 and MATLAB R2022b. The specific packages employed in each analysis are reported in Supplementary Materials 2-3.

#### Identifying distinct PSCI profiles

Latent class analysis (LCA) was used to identify common profiles of PSCI in a data-driven fashion. LCA assumes that the associations of the observed data arise from unobserved subgroups (i.e., classes) in the population. LCA is more powerful than k-means or hierarchical clustering (Cleland et al., 2000; Porcu & Giambona, 2017), as it enables comparisons of models of different numbers of classes^50^ and because it allows for class membership uncertainty (estimating the probability that an individual belongs to each identified class).

To establish the classes, only the binary impairment classifications on OCS subtests were included as observed variables.^51^ The LCA models used 10 repetitions (maximum of 50,000 iterations each) with different random starting values. The models were estimated including all partially available cases.^52,53^ To determine the ideal number of classes, models with n and n+1 classes were compared with the Bayesian Information Criterion (BIC), Akaike Information Criterion (AIC) and the adjusted Lo-Mendell-Rubin likelihood ratio test.^50,54,55,56^ A difference in the BIC and AIC larger than 2 is considered substantial evidence in favour of the more complex model.^57^ To investigate class distinguishability, the relative entropy (ranging from 0 = no separation to 1 = perfect separation) was calculated.^58^

#### Interpretation of PSCI profiles

To interpret LCA classes, the cognitive profiles of patients in each behavioural class were summarised. Class members were always compared to non-members to identify unique characteristics of each class. Specifically, each class’ probability of impairment and 95% confidence interval for each subtest was compared to that of patients belonging to other classes.

Neuroanatomical characteristics were compared between class members and non-members. First, we estimated differences in proportions of left- and right-hemispheric patients. Second, the ratio of lesion volume between class members and non-members was estimated. Third, differences in GCA and Fazekas scores were estimated. In addition, differences in proportions of younger vs. older stroke patients, education levels, acute versus chronic stroke and male patients were estimated. Model fits were satisfactory and all modelling details are reported in Supplementary Materials 2.

Mass-univariate voxel-based lesion analysis (VLSM) were conducted to evaluate whether classes were characterized by distinct lesion profiles. Each of these analyses included lesion volume as a covariate of no interest and employed permutation-based thresholds (2000 permutations) to correct for multiple comparisons (alpha = .05) (de Haan & Karnath, 2018). These analyses were performed with NiiStat^59^ and only considered voxels which were impacted in at least 10 patients (Supplemental Materials 3). Resultant significant voxels clusters (>10 contiguous voxels) were compared to anatomical atlases.

We also contrasted network disconnections of class members versus non-members, using a test that assumes that edges that differ between groups are clustered into components (i.e., a subnetwork of connected edges).^60^ This approach increases statistical power relative to mass-univariate statistical comparisons.^61,62^ First, we identified edges which were associated with class membership (uncorrected p<.01). Then, significant components of these edges were identified, estimating a family-wise p-value with 1000 permutations for the sum of edge weights of the component. As there are no clear guidelines on how edges should be thresholded^61,62^, we checked the differences in edge weights for significant versus non-significant edges (Supplementary Materials 3). Then, we quantified *network disconnection*. An edge was considered part of a functional network if at least one node belonged to the network (e.g. both inter- and intra-network connections) (Supplementary Materials 3). The VLSM and network analyses were only conducted for classes which contained at least 15 patients with available lesion data.

#### Does lesion topography drive PSCI profiles?

Finally, we evaluated the extent to which lesion anatomy drove the LCA solution using multivariate similarity analysis. This analysis determines to what extent patients who were behaviourally similar were also similar in terms of lesion anatomy. To quantify cognitive similarity, we computed the distance between the class probabilities for each pair of participants. We also computed neuro-anatomical (dis)similarity at the voxel, tract, and network level. Then we assessed the association between the cognitive and the neuroanatomical distances (Figure 1, Supplementary Materials 3).

**Figure 1.**
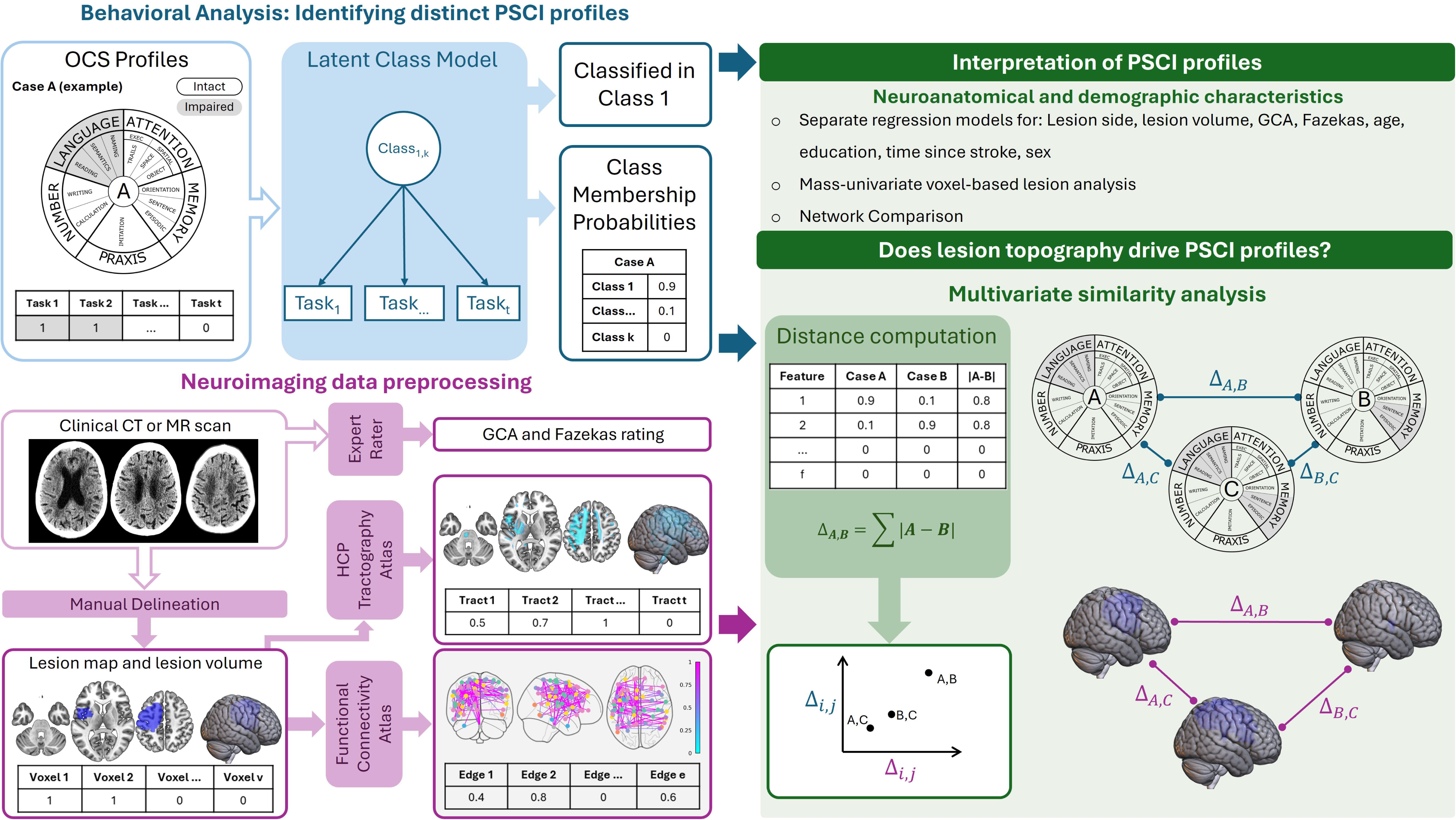
Overview of the analysis. Top left (blue) illustrates an OCS profile of a single case, the LCA model and output we obtain for case A from the LCA model. Bottom left (purple) illustrates preprocessing of clinical CT or MR scans. The right side (green) the association of cognitive profiles, neuroanatomy and demographic variables. For the multivariate similarity analysis, a distance is computed for each pair of cases (example of 3 cases) and the association of cognitive and neuroanatomical distances was then calculated.

The robustness of the similarity analysis was evaluated across key patient subgroups. For example, premorbid brain health decline may reduce the association between lesion location and the cognitive profile.^15^ In addition, haemorrhagic stroke patients can suffer from diffuse damage meaning that the relationship between cognition and lesion location could be weaker in this subgroup.^63^ Given that previous research has suggested that lesion location-behaviour relationships are strongest early after stroke onset, we also evaluated how this association depended on time after stroke.^36,63^ For all associations, we performed a leave-one-case-out sensitivity analysis which indicated robust results across subsamples.

## Results

### Participants

Patients were assessed on average 19 days post-stroke (*Mdn* = 7, *SD* = 35.7). A total of 43% of patients had a left-hemispheric stroke, 50% had a right-hemispheric stroke, and 7% had a bilateral stroke. Most patients had an ischemic stroke (81% ischemic, 19% haemorrhage) and 45% were female. The average age of the patients was 71 years (*SD* = 13.6). The average years of formal education was 11 (*SD* = 4). The lesion distribution and tract disconnections of patients with a lesion map are visualized in Figure 2.

**Figure 2.**
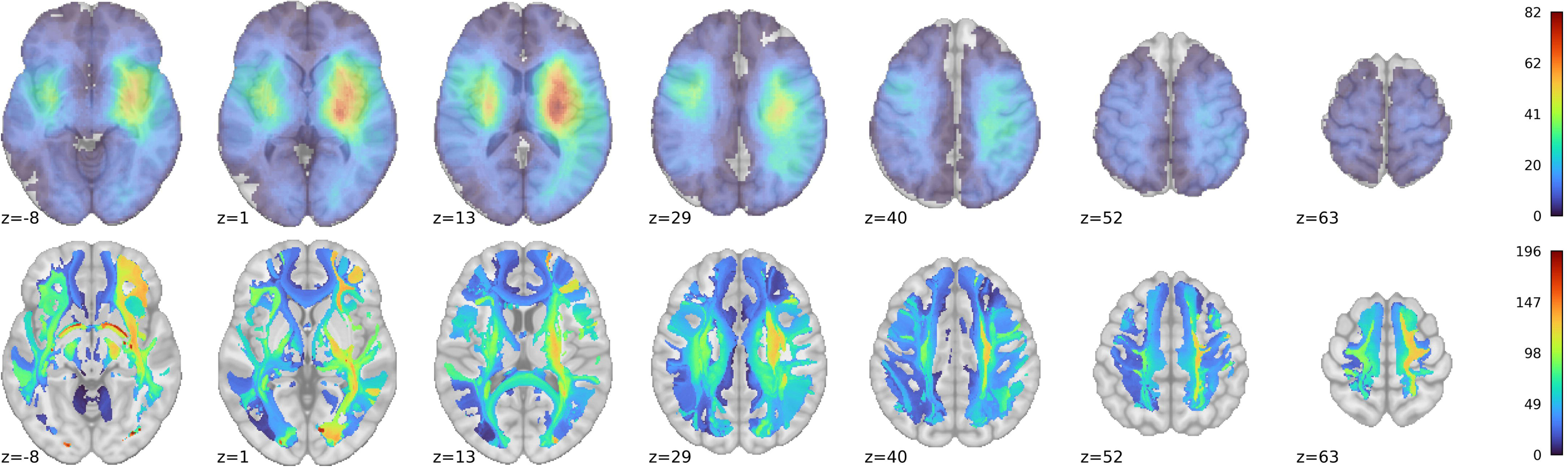
Lesion overlay and tract disconnections (n = 515). Number of patients with >= 50% disconnection in a tract. MNI slices -8 – 63 are presented in neurological convention (right hemisphere presented on the right side).

### PSCI can be distinguished into 5 or 13 profiles

Two candidate models of distinct cognitive profiles were retrieved: a 5-class model (best fitting according to the BIC index) and a 13-class model (best fitting according to the AIC and Likelihood Ratio Test) (Supplementary Materials 2). The relative entropy was .64, suggesting that there is a moderate level of class separability. The probability that an individual was a member of their respective class was on average 84% for the 5-class model (SD = 16) and 74% for the 13-class model (SD = 19). The probability that an individual was a member of another class was on average 4% for the 5-class model (SD = 7) and 2% for the 13-class model (SD = 4). The relationship between the two solutions is complex, with some classes mapping better onto each other than others across the solutions (Supplementary Figure S2.6). To interpret the models, we inspected the probability of impairment per OCS subtest for class members versus non-members and their neuroanatomical and demographic characteristics (Figure 3-6, Tables 1-2).

**Figure 3.**
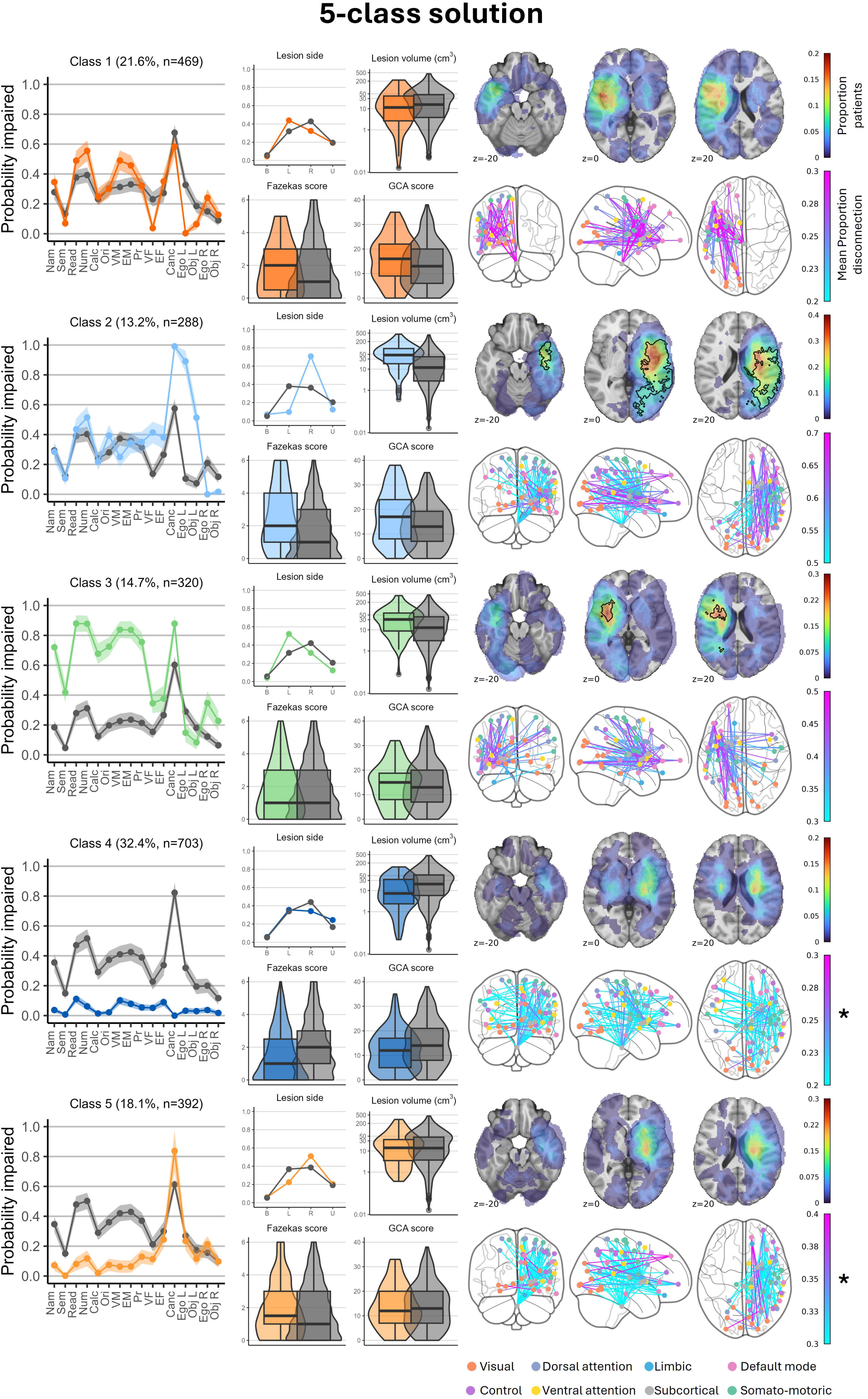
Characteristics of the 5-class model. Left panels: cognitive profiles and neuro-anatomical characteristics of class members (colour) versus others (grey). Right panel, top row: lesion overlay. A black contour indicates a region of significant voxels. Right panel, bottom row: 1% strongest and statistically significant network-level disconnections of class members. *= non-significant strongest disconnections depicted. Neuro-imaging is presented in neurological convention.

**Figure 4.**
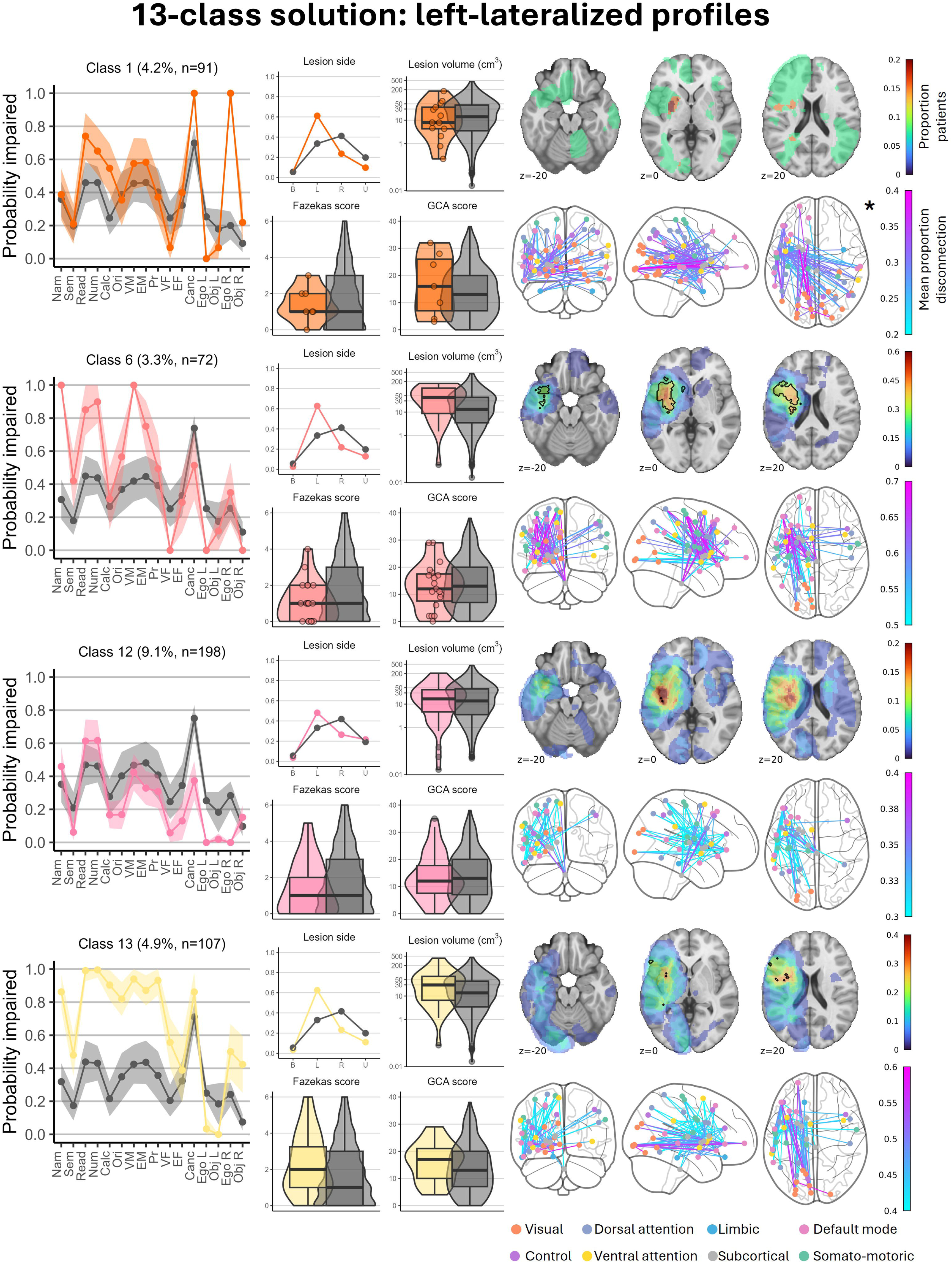
Characteristics of the left-lateralized profiles of the 13-class model. Left panels: cognitive profiles and neuro-anatomical characteristics of class members (colour) versus others (grey). Right panel, top row: lesion overlay (colours scaled such that red = 100% of class members with lesion map). A black contour indicates a region of significant voxels. Right panel, bottom row: 1% strongest and statistically significant network-level disconnections of class members. *= non-significant strongest disconnections depicted. Neuro-imaging is presented in neurological convention.

**Figure 5.**
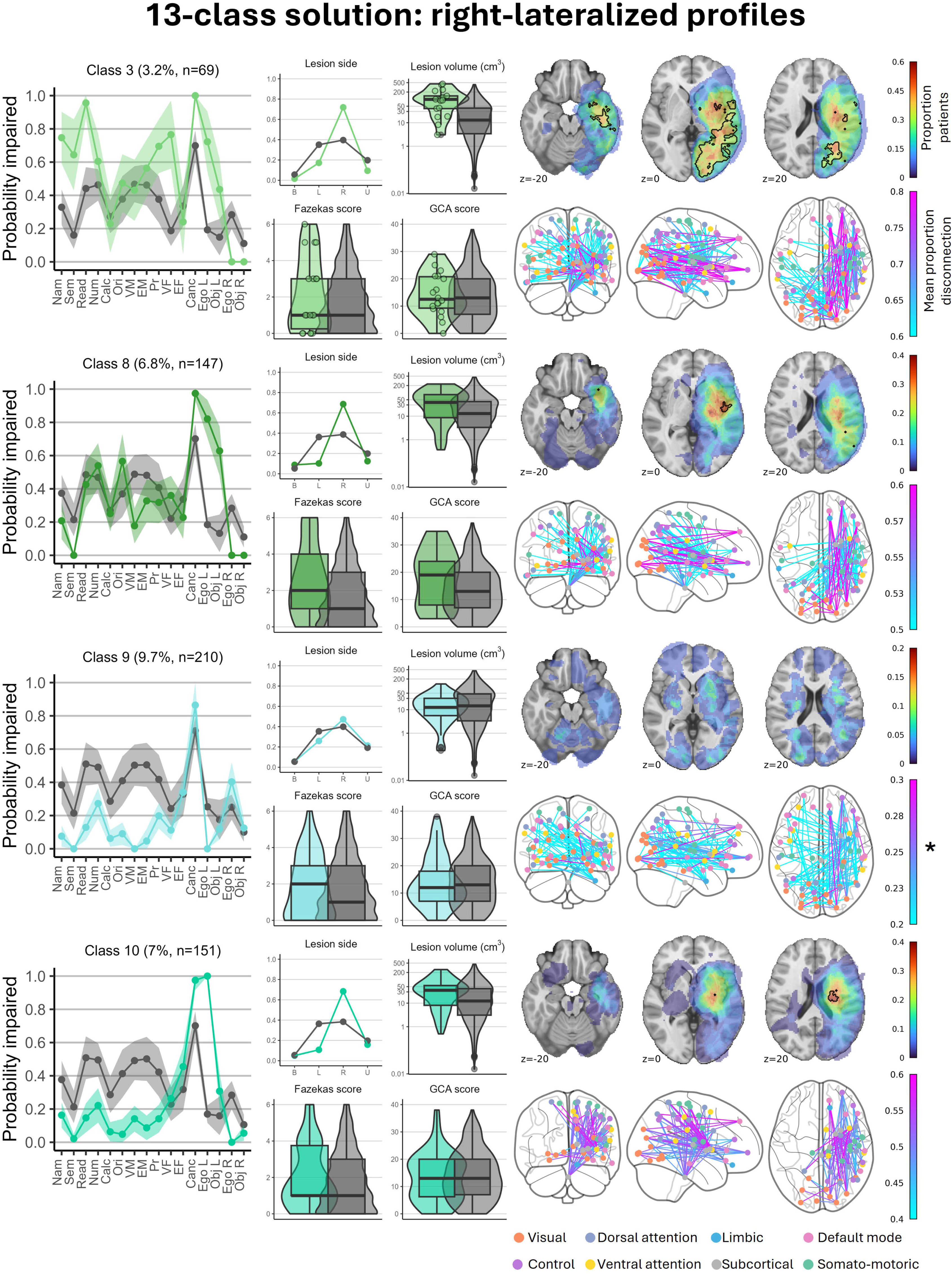
Characteristics of the right-lateralized profiles of the 13-class model. Left panels: cognitive profiles and neuro-anatomical characteristics of class members (colour) versus others (grey). Right panel, top row: lesion overlay. A black contour indicates a region of significant voxels. Right panel, bottom row: 1% strongest and statistically significant network-level disconnections of class members. *= non-significant strongest disconnections depicted. Neuro-imaging is presented in neurological convention.

**Figure 6.**
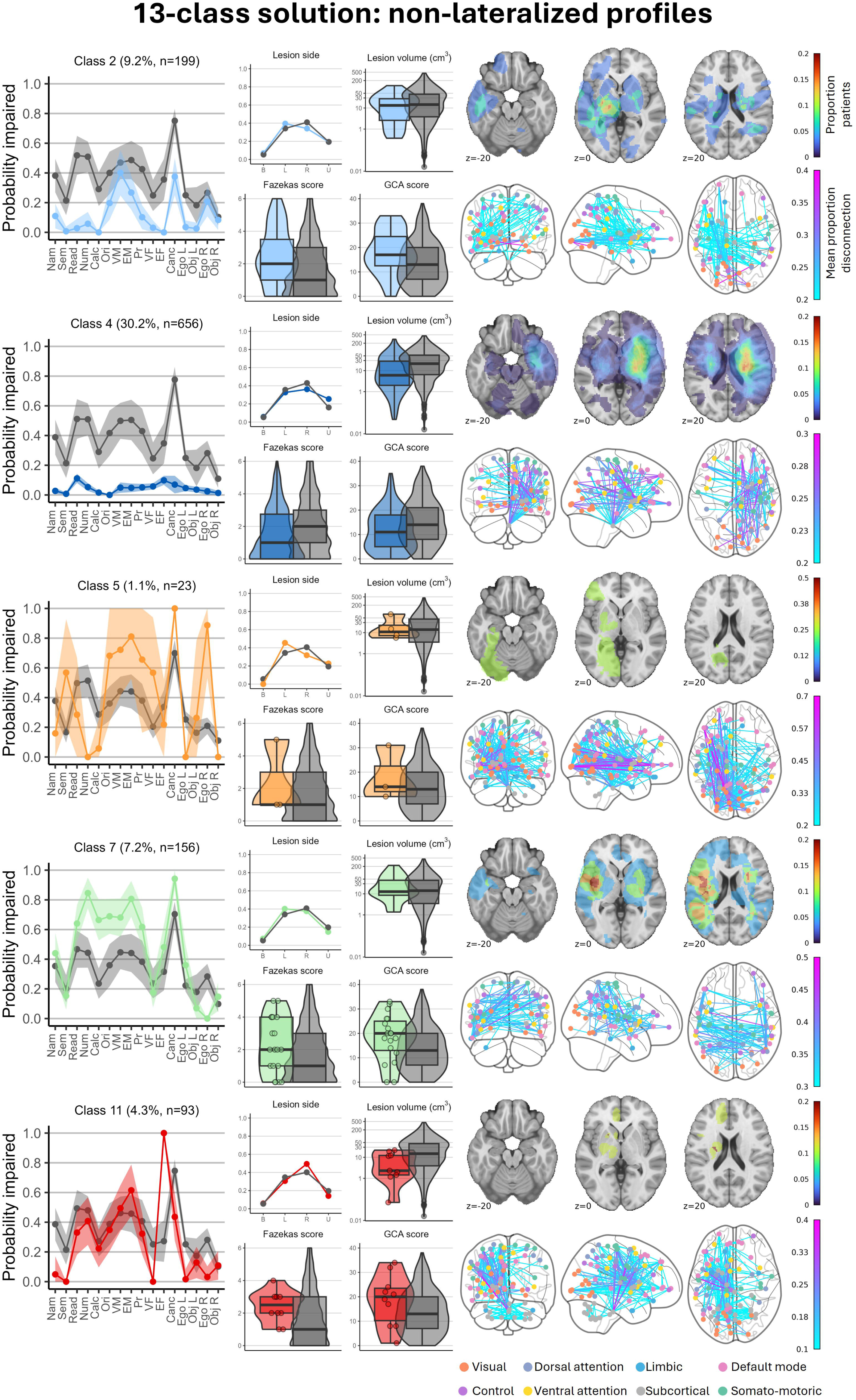
Characteristics of the non-lateralized profiles of the 13-class model. Left panels: cognitive profiles and neuro-anatomical characteristics of class members (colour) versus others (grey). Right panel, top row: lesion overlay. A black contour indicates a region of significant voxels. Right panel, bottom row: 1% strongest network-level disconnections of class members are depicted. Neuro-imaging is presented in neurological convention.

**Table 1.**
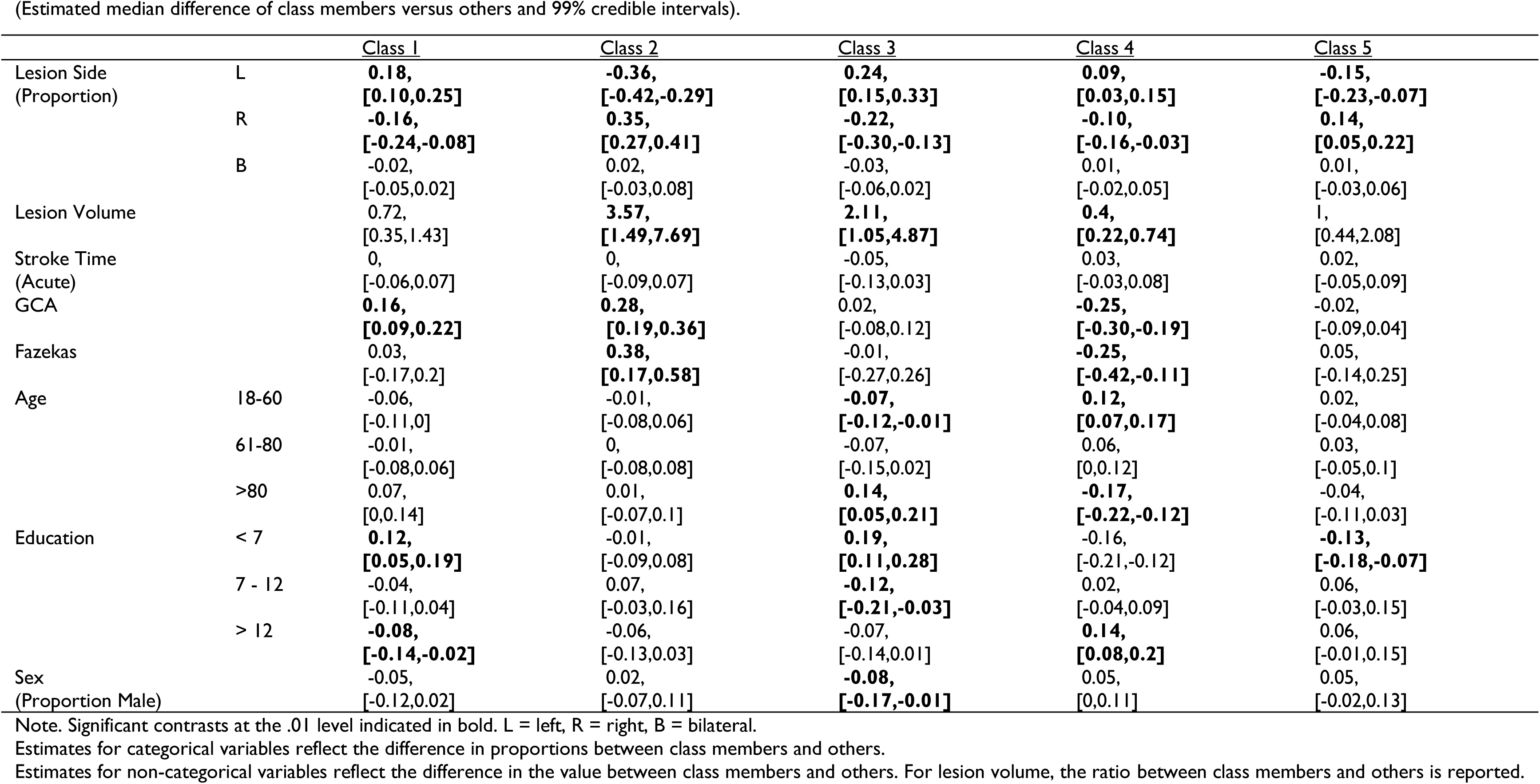
Characteristics of the 5-class solution. (Estimated median difference of class members versus others and 99% credible intervals).

**Table 2.**
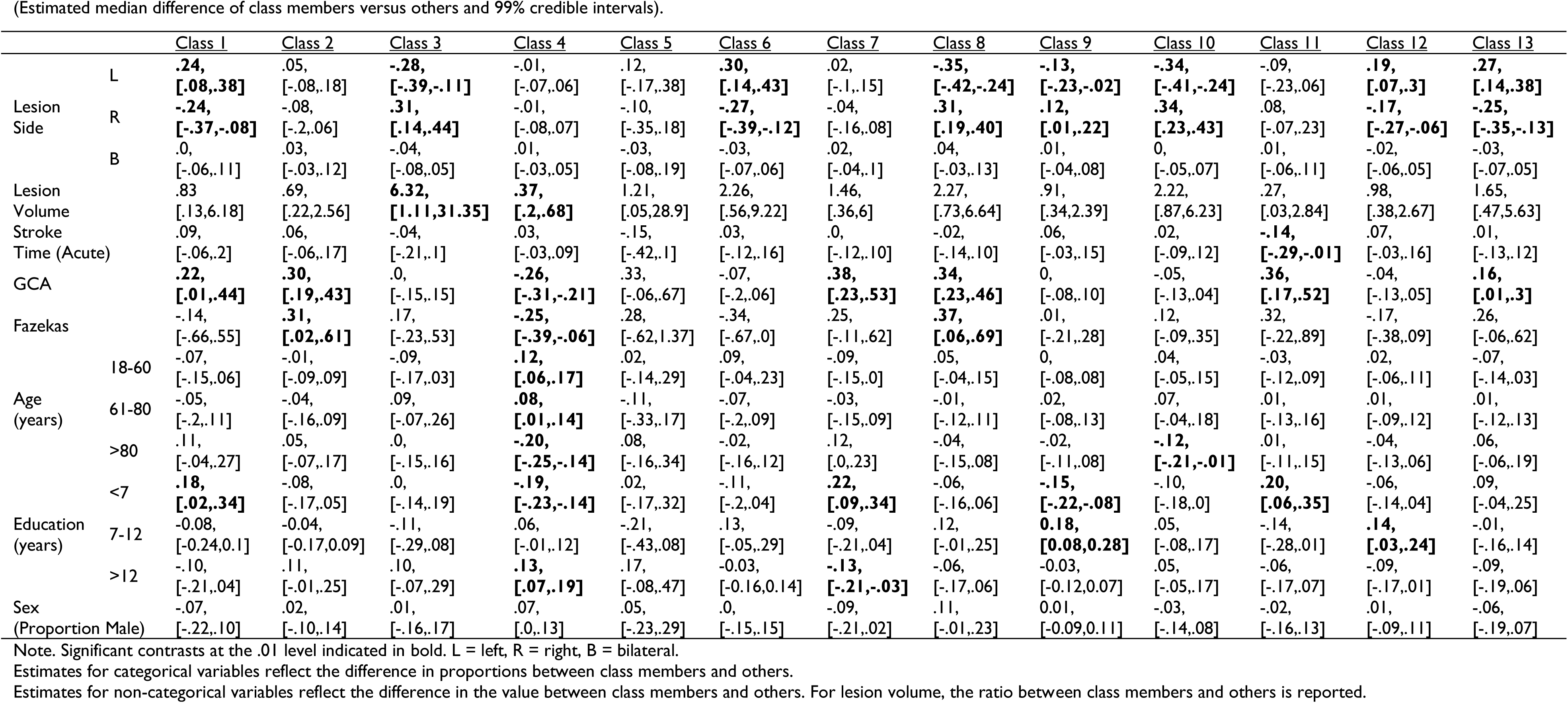
Characteristics of the 13-class solution. (Estimated median difference of class members versus others and 99% credible intervals).

### Interpretation of the 5 profiles

**Class 1 (“Memory”, n = 469)** was characterized by a comparably higher rate of numerical cognition and memory impairments coupled with an absence of visual field impairment and left egocentric neglect. Class 1 members were more likely left-than right-hemispheric stroke patients (E_LH_=.54, 99%CI [.48, .61], E_RH_=.40, 99%CI [.34,.47]) and had higher GCA ratings. Class 1 was associated with disconnection within 233 network edges spread across all networks but no significant voxel clusters. Class 1 members were more likely to be lower educated.

**Class 2 (“Left Neglect”, n = 288)** included primarily right hemisphere stroke patients (E_LH_=.11, 99%CI [.06, .16], E_RH_=.81, 99%CI [.73, .87]) with left egocentric and allocentric neglect. Class 2 members had comparably higher rates of visual field and executive function deficits. They were characterized by larger lesions and worse premorbid brain health. VLSM analysis identified two clusters of significant voxels associated with this class, impacting the right lateral occipital cortex, insular cortex, and angular gyrus (150 cm^3^, MNI = [46, -4, 2]) and the right middle frontal gyrus (0.13 cm^3^, MNI = [30, 14, 30]). Class 2 was associated with 773 disconnections, the largest portion of which were within the right-hemispheric visual network (18% of all visual connections).

**Class 3 (“Language”, n = 320)** was characterized by widespread impairment affecting language, numerical cognition, memory and praxis. Class 3 was associated with larger lesions and left-hemisphere stroke (E_LH_=.60, 99%CI [.52, .66], E_RH_ = .35, 99% CI [.29, .43]). Lesions in 4 voxel clusters in the left insula and putamen (16.3 cm^3^, MNI = [-32, 4, 8]), left precentral gyrus (0.35 cm^3^, MNI = [-46, 0, 42]), the left middle frontal gyrus (0.24 cm^3^, MNI = [-40, 24, 26]), and the left white matter (0.11 cm^3^, MNI = [-28, -60, 18]) were associated with Class 3. Class 3 was associated with 429 disconnections with the left-hemispheric default mode network most impacted (12% of all default mode connections). Class 3 members were also less likely younger and more likely lower educated.

**Class 4 (“No or mild impairment”, n = 703)** was characterized by a low probability of impairment. Class 4 members had smaller lesions, better premorbid health, were more likely younger and higher educated. Class 4 members were equally likely left- or right-hemispheric stroke patients (E_LH_ = .47, 99% CI [.42,.52], E_RH_ = .45, 99% CI [.40,.50]).

Finally, **Class 5 (“Attention”, n = 392)** was characterized by non-lateralised cancellation task impairment accompanied by low probabilities of impairment across all other subtests. Class 5 members were more likely right-hemisphere stroke patients (E_LH_ = .28, 99% CI [.22, .35], E_RH_ = .64, 99% CI [.57,.71]) and less likely lower educated.

There was no significant voxel cluster or network-level disconnection pattern associated with Class 4 or Class 5.

### Interpretation of the 13 profiles

#### Left-lateralized profiles

**Class 1 (“Right-sided neglect”, n = 91)** was characterized by right egocentric neglect and spared performance on the visual field test. In addition, Class 1 members exhibited a higher rate of sentence reading and numerical cognition impairment. Class 1 members were more likely left-hemispheric stroke patients (E_LH_ = .66, 99% CI [.50, .80], E_RH_ = .27, 99% CI [.14, .42]) with higher GCA ratings and a lower education level. There were insufficient lesion maps (n = 13) to investigate the association with lesion anatomy.

**Class 6 (“Global language & memory”, n = 72)** was characterized by impairments on language (picture naming and sentence reading), verbal memory and number writing. Class 6 had spared performance on the visual field test, and no left-sided egocentric neglect. Class 6 members were more likely left-hemispheric patients (E_LH_ = .71, 99% CI [.57,.84], E_RH_ = .24, 99% CI [.13, .38]). A cluster of significant voxels including the left insular cortex, putamen and inferior frontal gyrus (pars opercularis) (volume = 28.8 cm^3^, peak z-score at MNI = [-30, 6, 12]), and a second cluster impacting the left insular cortex and left central opercular cortex (0.30 cm^3^, MNI = [-30, -20, 26]) were associated with Class 6. Class 6 was characterized by 360 disconnections in several networks, among which the left default mode network was most disconnected (12%).

**Class 12 (“Expressive language”, n = 198)** was characterized by a low probability of impairment across all tasks, except for sentence reading, number writing and picture naming, with mostly intact memory performance compared to class 6. Class 12 membership was associated with left-hemispheric stroke (E_LH_ = .62, 99% CI [.51, .72], E_RH_ = .33, 99% CI [.23, .43]) and two voxel clusters, one centred in the left Heschl’s Gyrus (0.19 cm^3^, MNI = [-42, -20, 8]) and the second within the left planum polare (0.18 cm^3^, MNI = [-44, -10, -6]). Class 12 was characterized by 186 disconnections, with the left-hemispheric default mode network being the most impacted (7%). Class 12 patients were more likely part of the middle education level than others.

**Class 13 (“Severe language & neglect”, n = 107)** was characterized by high probabilities of impairment across language, numerical cognition, memory, praxis, visual field impairments, and right-sided neglect. Class 13 included mainly left-hemispheric stroke patients (E_LH_ = .69, 99% CI [.58, .80], E_RH_ = .26, 99% CI [.16, .37]) with higher GCA ratings. Class 13 was associated with 7 left-hemispheric voxel clusters: precentral gyrus (0.55 cm^3^, MNI = [-34, 2, 22]), inferior frontal gyrus (pars triangularis) (0.34 cm^3^, MNI = [-58, 30, 10]), insula (0.14 cm^3^, MNI = [-36, 6, -4]), hippocampus (0.12 cm^3^, MNI = [-36, -22, -12]), inferior frontal gyrus (pars opercularis) (0.10 cm^3^, MNI = [-58, 20, 24]), and two clusters in the white matter (0.96 cm^3^, MNI = [-40, -40, 2]; 0.88 cm^3^, MNI = [-40, -36, -6]). Class 13 was characterized by 266 disconnections in many networks, among which the left dorsal attention network was most disconnected (8%).

#### Right-lateralized profiles

**Class 3 (“Severe left-sided visuospatial impairment”, n = 69)** mainly included right-hemispheric stroke patients (E_LH_ = .19, 99% CI [.08, .33], E_RH_ = .78, 99% CI [.62, .90]) who had impairments on tests involving a visual component. Class 3 was associated with larger lesions, and 9 significant voxel clusters mainly impacting the right visual cortex. The largest of these voxel clusters (70.14 cm^3^ volume) impacted the lateral occipital cortex (inferior division), middle temporal gyrus (posterior division) and the intracalcarine cortex (MNI = [24,-80,6]). The remaining voxel clusters were centred in the right middle frontal/precentral gyri (0.86 cm^3^, MNI=[32,-6,26]), amygdala (0.32 cm^3^, MNI= [20, -6, -10]), precentral gyrus (0.29 cm^3^, MNI=[56, 4, 32], supramarginal gyrus (0.29 cm^3^, MNI= [66, -36, 30]), inferior frontal gyrus pars opercularis (0.27 cm^3^, MNI=[60, 16, 6]), insular cortex (0.15 cm^3^, MNI = [32, -2, 14]), hippocampus (0.14 cm^3^, MNI=[30, -18, -16]), and putamen (0.13 cm^3^, MNI=[26, -4, 10]). Class 3 was associated with 615 network-level disconnections with the right visual network being the most impacted (18%).

**Class 8 (“Severe left-sided neglect”, n = 147)** was characterized by left egocentric and allocentric neglect, coupled with lower probabilities of impairment on picture naming and verbal recall tasks. Class 8 patients primarily had right hemisphere lesions (E_LH_ = .12, 99% CI [.06, .21], E_RH_ = .77, 99% CI [.67, .86]) and worse premorbid brain health. Class 8 was associated with a significant cluster of voxels in the right insular cortex and superior temporal gyrus (0.62 cm^3^, MNI = [48, -10, 0]). Class 8 was associated with 476 disconnections among which the right visual network was most disconnected (12%).

**Class 10 (“Left-sided egocentric neglect”, n = 151)** was characterized by left-sided egocentric neglect coupled with low impairment rates on other tasks mostly due to right-hemispheric stroke (E_LH_ = .13, 99% CI [.07, .22], E_RH_ = .80, 99% CI [.70, .88]). Class 10 was associated with significant voxel clusters in the right putamen, thalamus and white matter (3.88 cm^3^, MNI = [22, -18, 18]), right precentral gyrus (0.08 cm^3^, MNI = [44, -10, 32]), and postcentral gyrus (0.08 cm^3^, MNI = [44, -16, 32]). Class 10 members had 493 significant disconnections among which the ventral attention network in the right hemisphere was most disconnected (11%).

**Class 9 (“Attention”, n = 210)** was characterized by non-lateralised impairment on the cancellation test cooccurring with low impairment rates across other tests. The majority of patients had a right-hemispheric stroke (E_LH_ = .33, 99% CI [.24,.43], E_RH_ = .60, 99% CI [.49, .69]) and Class 9 members were less likely lower educated. Class 9 was not significantly associated with any voxels nor network-level disconnections.

#### Non-lateralized profiles

Finally, there were 5 non-lateralized profiles (Figure 6) which were not significantly associated with lesion side, location and network disconnections.

**Class 2 (“Premorbid cognitive impairment”, n = 199)** was characterized by moderate impairment probabilities on memory tasks or the cancellation task. Notably, Class 2 members had worse premorbid brain health. Class 2 was not included in VLSM, as there was insufficient lesion overlap.

**Class 4 (“No or mild impairment”, n = 656)** was characterized by a low probability of impairment across all subtests. Class 4 members had significantly smaller lesions, better premorbid brain health, more likely to be younger and higher educated.

**Class 5 (“Mild right-sided neglect”, n = 23)** was characterized by right-sided egocentric neglect. However, Class 5 consisted of few patients and therefore had a high uncertainty regarding the impairment probabilities and associated covariates.

**Class 7 (“Low cognitive reserve”, n = 156)** was characterized by higher probabilities of impairment within the numerical and memory domains. Class 7 patients had higher GCA ratings and were more likely lower educated. There was insufficient lesion overlap to examine the relation with lesion location.

**Class 11 (“Executive impairment”, n = 93)** was characterized by executive function impairment which occurred in the absence of neglect and visual field impairments. Class 11 patients had higher GCA ratings, were less likely acute stroke patients and more likely lower educated. There were not enough lesion maps (n = 9) available to assess Class 11’s relation with lesion location.

### Lesion neuroanatomy does not drive PSCI profiles

Last, we evaluated the extent to which the cognitive profiles as identified by the LCA model were driven by lesion neuroanatomy (Figure 1). First, cognitive and neuroanatomical similarity was evaluated for all patients with a lesion map. Within this group, all lesion metrics were significantly positively associated with cognitive similarity (Figure 7). Associations of network-level disconnectivity (r(514^1^) = .16, 95%CI = [.15, .17]) and lesion volume (r(514) = .15, 95%CI [.14,.15]) were highest, followed by tract disconnections (r(514) = .11, 95%CI = [.11, .12]) and lesion location (r(514) = .04, 95%CI = [.03, .04]). However, these associations were small (< .20)^64^ indicating that the similarity in cognitive profiles was only partially explained by similarity in lesions and their corresponding disconnections.

**Figure 7.**
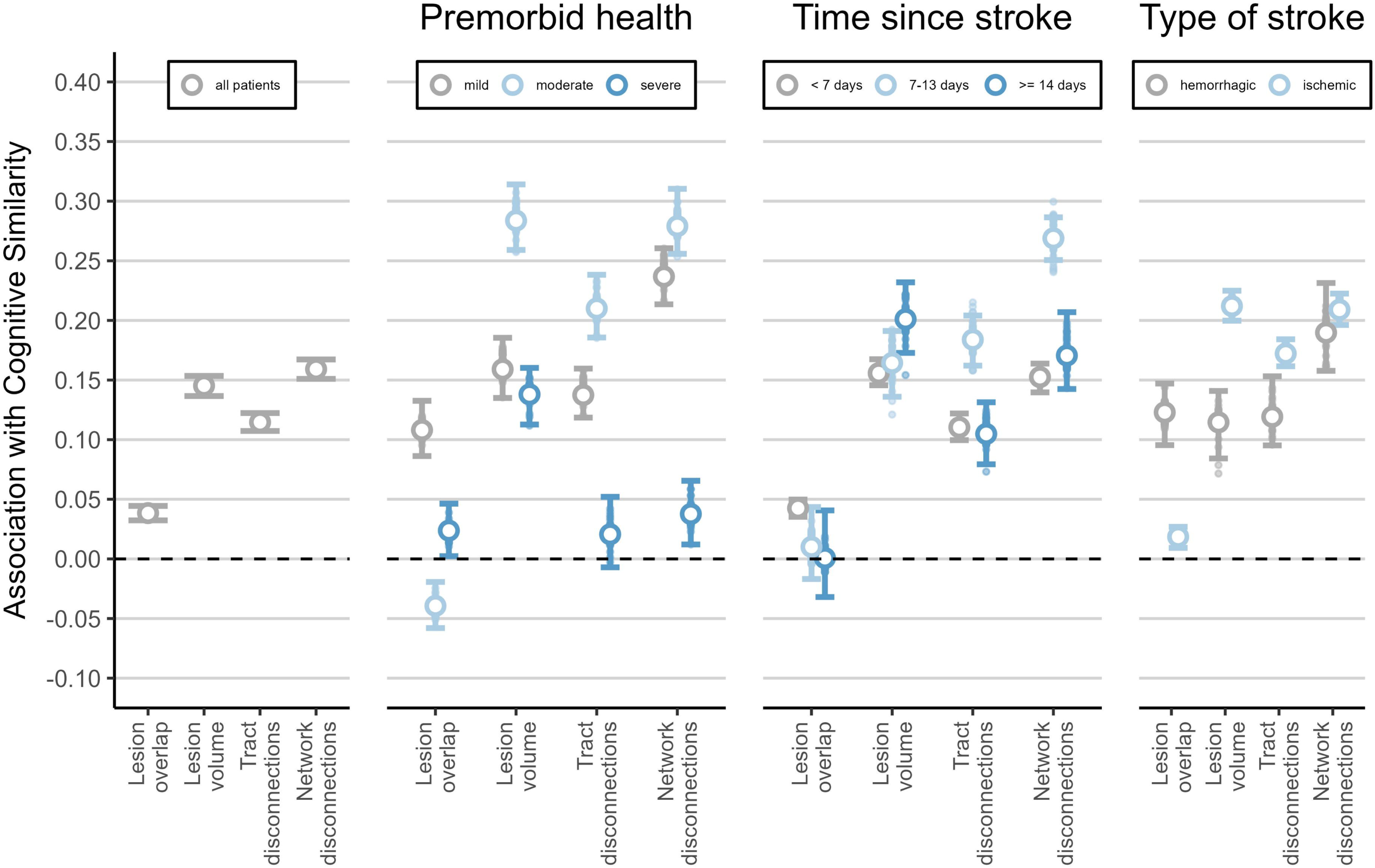
Associations of cognitive and neuro-anatomical similarity (lesion overlap, lesion volume, tract- and network disconnections). The error bars represent the 95% confidence intervals of the correlation. The small dots outside the error bars represent the extreme correlations based on a leave-one-out procedure to assess influential observations.

Next, analyses were conducted to evaluate whether these results were modulated by premorbid brain health (i.e. Global Cortical Atrophy Score + Fazekas score), the type of stroke damage (e.g. haemorrhages vs. ischemic stroke), and the time between stroke and testing. Within premorbid brain health analyses, the association between cognition and network-level disconnections was moderate in the two groups with less severe atrophy and white matter lesions (mild: r(94) = .24, 95%CI [.21, .26], moderate: r(89) = .28, 95%CI [.26, .31]), and was very low in the severe group (r(92) = .04, 95%CI [.01, .07]) (Figure 4). Lesion location had the highest association with cognitive similarity in the mild brain health group (r(94) = .11, 95%CI [.08, .13]) and this association was low in the severe brain health group (r(92) = .02, 95%CI [.00, .05]). Lesion volume had a similar association with cognitive similarity in the mild brain health (r(94) = .16, 95%CI = [.14, .19]) and severe brain health group (r(92) = .14, 95%CI [.11, .16]). These results suggest a stronger association between lesion location (and corresponding tract and network-level disconnections) with the cognitive profile in patients with less severe atrophy and white matter lesions. In the group of patients with more severe atrophy and white matter lesions, lesion volume was the best predictor of cognitive profiles, suggesting that severity of impairments may become more important than the type of impairment.

For time since stroke, the association of cognitive similarity and disconnection profile was similar between the hyper-acute stroke patients (< 7 days) (r(344) = .15, 95%CI = [.14,.16]) and patients tested later after stroke (r(59) = .17, 95%CI = [.14, .21]). The group tested in between 1- and 2-weeks post-stroke had the highest association (r(66) = .27, 95%CI = [.25, .29]). As time after stroke increased, the association with lesion volume increased, progressing from .16 (95%CI = [.15,.17]) to .20 (95%CI = [.17,.23]), while the association with lesion location decreased, going from .04 (95%CI = [.04,.05]) to .00 (95%CI = [-.03, .04]).

Last, we assessed the associations between cognitive and neuroanatomical similarity in haemorrhagic versus ischemic stroke patients. The disconnection profile had a similar association with cognition in both groups (haemorrhage: r(78) = .19, 95%CI = [.16,.23], ischemic: r(257) = .21, 95%CI [.20,.22]). Lesion volume was more strongly associated with cognition in the ischemic (r(257) = .21, 95%CI [.20,.22]) than haemorrhagic group (r(78) = .11, 95% CI [.08,.14]), while lesion location was less strongly associated with cognition in the ischemic (r(257) = .02, 95%CI = [.00, .03]) than haemorrhagic group (r(78) = .12, 95%CI [.09,.15]).

## Discussion

This study’s results indicate that patterns of PSCI can be captured by underlying behavioural classes, and that these classes cannot be entirely explained by differences in lesion anatomy. Overall, these results provide novel insight into the underlying structure of PSCI impairment, providing important theoretical groundwork necessary to support future translational work.

This investigation yielded two viable distinctions of PSCI profiles: a 5- and a 13-class solution. Some aspects of the simpler, 5-class solution are comparable to the PCA solutions reported by Corbetta et al. (2015) and Bisogno et al. (2021), but there are also key differences. That is, our 5-class solution captures 2 profiles of classic cognitive deficits which occur following left and right-lateralised stroke (left stroke with aphasia, right stroke with neglect). However, our 5-class solution also captures profiles that do not represent such classical deficits. For example, class 1 included patients with non-language impairments (e.g. memory, numerical cognition) while class 5 captured right hemisphere (and some left hemisphere) patients with non-lateralised attention deficits without neglect. Importantly, our 5-class solution also reflects that not all stroke survivors exhibit severe stroke-related cognitive impairment (Class 4). This class was associated with smaller lesions, better premorbid health, younger age and higher education levels, highlighting the protective role of brain and cognitive reserve.^6,8,16,17,65^

The 13-class solution provided a richer perspective on PSCI, distinguishing 4 left-lateralized, 4 right-lateralized and 5 non-lateralized profiles. These non-lateralised profiles capture important variability in PSCI profiles, particularly with respect to premorbid cognitive status.

In the complex solution, the right-lateralised profiles were characterized by different types of visual-attentional impairments. Specifically, class 3 captured severe and global neglect and visual field impairment which impacted performance on all tasks involving a visual component. This profile was linked to large lesions impacting regions within (and connections between) early visual areas and regions of the posterior parietal cortex traditionally associated with neglect.^66^ This captures the common clinical presentation of comorbid neglect and visual field impairment. A second group (class 8) exhibited left egocentric and allocentric neglect and was associated with lesions in the right insula and superior temporal gyrus (both regions associated with neglect^66–68^). Interestingly, this group exhibited worse premorbid brain health compared to other groups. This result aligns with previous work suggesting that older patients with worse brain reserve were more likely to have spatial neglect^65^ and adds to this that patients with worse premorbid brain health have a higher risk of presenting multiple comorbid, rather than isolated neglect deficits.

The third right-hemisphere class (Class 10) captured cases of left egocentric neglect which occurred with few comorbidities, and was linked to lesions in the right putamen, thalamus and pre- and postcentral gyri, which have previously been associated with directional motor biases and egocentric neglect.^69,70^ Interestingly, this class was mainly linked to disconnection in the right ventral attention network, while the other neglect classes were mainly linked to visual network disconnection. Taken together, the three left-neglect classes align with previous conceptualisations of neglect as a deficit which represents a common symptom of multiple underlying causes.^71,72^ The last right-hemisphere class (class 9) included patients with non-lateralised attentional impairment. This class is analogous to class 5 from the 5-class model as it similarly includes patients which likely suffer from general attentional deficits.

In terms of left-hemisphere profiles, two classes captured patients with differing severity of aphasia. Class 6 represented the classical, pure aphasia profile (anomia, alexia and agraphia) and was accordingly associated with large lesions affecting key language areas.^73–75^ Class 13 included patients with aphasia occurring alongside widespread multi-domain impairments. In line with past work, this globally impaired group exhibited worse premorbid brain health ^8,16^ and was associated with large lesions affecting language and memory regions.^76^ Notably, these two profiles do not align with classic aphasia distinctions.^24,77^ This finding suggests that subtypes which are often prioritised in the neuropsychological literature may capture theoretically important special cases rather than representing the symptom variability characteristic of the clinical population.

The third left-hemisphere group (Class 1) was characterised by right-lateralised neglect. We have previously found interhemispheric disconnections to be associated with right neglect^78^, and the present study expands on this with interhemispheric disconnections mainly between left frontotemporal areas and right posterior parietal regions. The remaining group (Class 12) included patients with left hemisphere strokes and comparatively mild rates of language and numerical cognition impairment.

The remaining 5 cognitive profiles were not lateralised and were not associated with any significant lesion correlates, but reflect the importance of premorbid brain health. First, Class 4 was characterised by low rates of cognitive impairments coupled with better premorbid brain health, younger age, and higher education levels (analogous to Class 4 from the 5-class solution). Class 2 was characterised by memory impairments and non-lateralised cancellation task impairment, likely capturing pre-morbid cognitive decline.^22^ Class 11 included patients with executive function impairment. Given that this class exhibited lower lesion sizes coupled with worse atrophy and white matter integrity, it is likely that executive dysfunction is more closely related to general brain health than to the acute stroke event. Class 5 captured a small portion of patients who exhibited right-lateralised neglect coupled with memory and praxis impairments. Finally, class 7 captured patients with numerical cognition and memory impairment who exhibited lower education levels and worse atrophy. These results highlight that the clinical picture of premorbid impairment and cognitive reserve can be distinctly and qualitatively different from classical PSCI profiles which are more linked to lateralised lesions.

Although several profiles mapped onto lesion locations, lesion location itself played a limited role in explaining profiles, as lesion similarity was only weakly associated with cognitive similarity. Our results highlight key factors which limit the explanatory power of lesion location. Mainly, lesion location was less predictive of cognitive similarity for patients with worse premorbid health. This finding aligns with past work suggesting that the impact of stroke lesions may be modulated by general brain health.^7,15^ In patients with severe atrophy, lesion volume was the strongest determinant of PSCI profile. This suggests that in populations with poor premorbid health, stroke severity may play a stronger role in determining cognitive consequences than lesion location.

Additionally, lesion anatomy was a better predictor of the PSCI profile in patients assessed in the hyper-acute phase. This result is likely driven by cognitive recovery occurring within the very early period post-stroke, which makes brain-behaviour relationships less clearly defined as time progresses following stroke.^36,79^ Notably, disconnection patterns were identified as an important driver of cognitive variability as lesion-induced disconnections at the tract- and network level were stronger predictors of the PSCI profile than lesion location itself. This finding is in line with past studies illustrating that disconnection metrics help account for important variability in post-stroke brain behaviour relationships.^44,80^

This study employed routinely collected data including a short cognitive screen and clinical neuroimaging. While this approach maximises the size and representativeness of this study, it is possible that this approach may not fully capture the association between lesion and cognitive profiles. For example, more extensive neuropsychological batteries could be used to tease apart more fine-grained relationships.^80,81^ In addition, in-vivo tractography could be used to capture key sources of variability in disconnectivity which may account for a significant portion of the variance in cognitive profiles.^15,76^ Additionally, the Fazekas ratings on the clinical CT scans, are likely underestimating the importance of white matter integrity. Importantly also, individual tests may not directly map onto a single cognitive function. Latent class models cannot distinguish between this case and true comorbidities, meaning that they can overestimate the number of subpopulations.^82^ For this reason, it remains important for future work to explore whether more advanced models can provide a better account of the structure of PSCI, such as hybrids of factor and latent class models.^82^ Moreover, future studies must investigate the replicability of the PSCI profiles and investigate whether the PSCI profiles predict differential recovery.^83^

Overall, the results of this study reveal that PSCI is heterogeneous, encompassing both domain-specific profiles linked to focal lesion sites and profiles that are more strongly associated to premorbid health and demographic factors. Focusing merely on low-dimensional solutions of PSCI may reveal the strongest factors, but more complex solutions may help capture critical cognitive profiles present in real-world clinical populations. Future studies can aim to build on this work by exploring whether cognitive profiles can be used to inform clinical care.

### Data availability

The behavioural data of the UK-OCS are publicly available on the DPUK platform. The OCS-NL data and main scripts to replicate our analysis are available on https://figshare.com/account/articles/28387994. The lesion masks can be made available upon request.

## Footnotes

1 The number of patients on which the association was based.

