## Supplementary Materials 1 for "Post Stroke Cognitive Impairment: more than a lesion-symptom model"

### Supplementary Materials 1. Description dataset

#### Patient recruitment

Patients were recruited in several studies with various selection criteria from study to study (Table S1.1). In the UK studies patients were either recruited within 3 weeks or 10 weeks post-stroke. In Italy, patients were included from 72 hours until 6 months post-stroke. In contrast, in Belgium there were no explicit criteria regarding time post-stroke. All studies recruited adult stroke patients. In the Italian study, only first-ever stroke patients were recruited, while in the UK and Belgian studies this was not a requirement to participate. Studies recruited patients at acute stroke units and rehabilitation centers.

**Table S1.1.** Information about patient recruitment

| Country | Reference | N | Hospital | Time since stroke | Age range (years) | Study-specific In- and exclusion criteria |
|---|---|---|---|---|---|---|
| UK | (Demeyere et al., 2015) | 208 | Acute stroke unit Oxford | ≤ 3 weeks | >=18 (no maximum) | / |
| UK | (Demeyere et al., 2016) | 200 | Acute stroke unit Oxford | ≤ 3 weeks | Not reported | / |
| UK | (Demeyere et al., 2019) | 821 | 37 sites | ≤ 10 weeks | 18 - 90 | Able to concentrate for 1 hour. Have sufficient language comprehension to pass first OCS tests. |
| Italy | (Mancuso et al., 2018) | 325 | 14 rehabilitation centers | ≥72 hours to ≤ 6 months | 18 - 90 | First-ever stroke. No premorbid psychiatric or neurological disease. |
| Belgium | (Huygelier et al., 2022) | 197 | 3 acute stroke, 3 rehabilitation units | ≤ 6 months | >=18 (no maximum) | / |

The demographic characteristics of stroke patients varied from country to country (Table S1.2). Belgian stroke patients were slightly younger than UK and Italian stroke patients on average.

**Table S1.2**. Patient characteristics by country.

| | | UK (n = 1206) | IT (n = 684) | BE (n = 282) |
|---|---|---|---|---|
| Age | M | 72.4 | 71.1 | 64.8 |
| | Mdn | 75 | 73 | 67 |

| | | | | |
|---|---|---|---|---|
| | SD | 13.6 | 12.8 | 14 |
| | Min-Max | 18-98 | 24-96 | 21-91 |
| Years of education | M | 11.7 | 8.6 | 12.4 |
| | Mdn | 11 | 8 | 12 |
| | SD | 2.9 | 4.5 | 3.6 |
| | Min-Max | 5-30 | 1-25 | 5-25 |
| Sex | % Female | 45% | 45% | 39% |
| Days since stroke | M | 8.7 | 37.6 | 22.7 |
| | Mdn | 4 | 20 | 15 |
| | SD | 14.4 | 54.7 | 24.4 |
| | Min-Max | 0-208 | 0-567 | 0-182 |
| Stroke type | % Ischemic | 84% | 77% | 81% |
| Lesion side | % Left | 46% | 40% | 38% |
| | % Right | 48% | 56% | 44% |
| | % Bilateral | 5% | 4% | 18% |

#### Behavioural data: missing data

A total of 577 patients did not complete all OCS subtests. For these patients, the average number of missing subtests was 2.5 (Mdn = 1, SD = 2.6). Only 60 participants had missing data on more than 6 of the 12 subtests (Figure S1.1A). The Little MCAR test demonstrated that missing data was not completely random (*MCAR(2525) = 5100, p < .01, 226 missing patterns*). Missing data occurred more frequently for certain subtests, such as the executive function and cancellation test (Figure S1.1B).

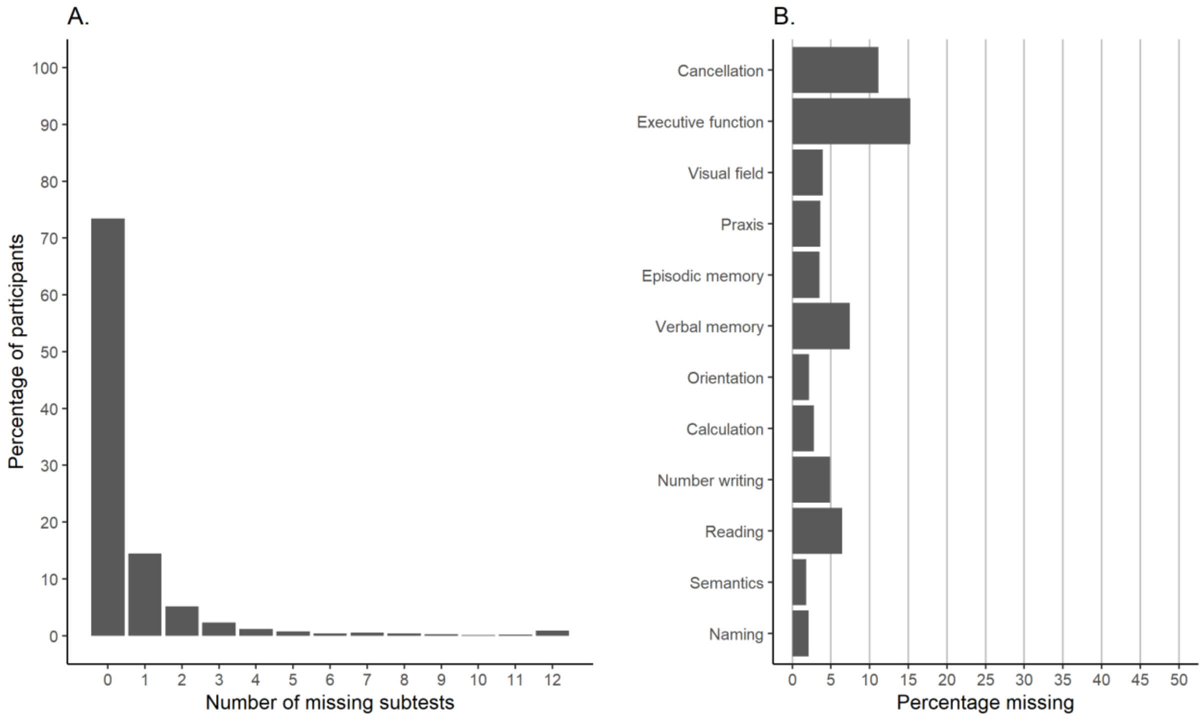


*Figure S1.1. Distribution of the number of missing subtests (A) and the percentage missing observations per OCS subtest (B).*

There was evidence for a significant difference between patients who completed and did not complete the OCS in age and years of formal education (Table S1.3). Completion rates depended on country with 90% of patients completing all subtests in the Belgian dataset, while this was 71% for the UK and 70% for the Italian dataset. In addition, patients with ischemic stroke were more likely to complete the OCS (76.5%) than patients with a haemorrhagic stroke (69%). According to the Bayes Factor (BF), there was no association of OCS completion with lesion side and the relation with sex was unclear (Table S1.3).

**Table S1.3. Patient characteristics associated with incomplete OCS data.**

| | Complete OCS | Incomplete OCS | Statistical test | Result |
|---|---|---|---|---|
| Age (years) | M = 70, Mdn = 72, SD = 13.5 | M = 73.6, Mdn = 77, SD = 13.7 | Welch two-sample t-test | t(912.92) = -5.21, p < .001, 95% CI = [-4.9, -2.2], $BF_{10}$ = 44614 |
| Education (years) | M = 10.91 Mdn = 11 SD = 3.9 | M = 9.96 Mdn = 10 SD = 4.1 | | t(637.83) = 4.19, p < .001, 95% CI [0.51, 1.40], $BF_{10}$ = 548.4 |
| Country (UK, IT, BE) | n = (859, 482, 254) | n = (347, 202, 28) | Bayesian Contingency Table Test | $BF_{10}$ > 5000 |
| Type of stroke (Ischemic, Haemorrhage, unknown) | n = (1076, 232, 287) | n = (331, 104, 142) | | $BF_{10}$ = 68 |
| Lesion side (Left, Right, Bilateral) | n = (531, 661, 93) | n = (219, 220, 26) | | $BF_{10}$ = 0.06 |
| Sex (Female, Male) | n = (691, 902) | n = (273, 292) | | $BF_{10}$ = 0.55 |

### Supplementary Materials 2. Behavioural analysis

#### Model comparison

The LCA models were fit with the poLCA R package (Linzer & Lewis, 2011).

To determine the optimal number of classes, different LCA models are contrasted with increasing number of classes until the model no longer converges. For each contrast, the difference in the AIC and BIC indices was computed (Table S2.1). In addition, the Likelihood Ratio Test was performed (Table S2.1). The AIC index and Likelihood Ratio Test indicated a 13-class model as the best fitting model, while the BIC indicated the 5-class model as the best fitting model. The relative entropy for each number of classes decreases from a model with two classes to a model with five classes and remained relatively stable after this point (Figure S2.1). The probability that a participant belongs to a class for the 5- and 13-class solution are depicted in Figure S2.2, for the class to which the participant was assigned to and the other classes.

**Table S2.1. Model comparison**

| M0 (k) | M1 (k+1) | Δ AIC | Δ BIC | LMR (17) | P-value |
|---|---|---|---|---|---|
| 1 | 2 | -3532.94 | -3436.32 | 3412.53 | < .001 |
| 2 | 3 | -678.17 | -581.55 | 681.34 | < .001 |
| 3 | 4 | -301.37 | -204.76 | 320.86 | < .001 |
| 4 | 5 | -107.25 | -10.64 | 135.14 | < .001 |
| **5** | 6 | -70.28 | 26.34 | 99.76 | < .001 |
| 6 | 7 | -52.17 | 44.45 | 82.44 | < .001 |
| 7 | 8 | -34.67 | 61.95 | 65.70 | < .001 |
| 8 | 9 | -22.48 | 74.14 | 54.04 | < .001 |
| 9 | 10 | -14.08 | 82.54 | 46.00 | < .001 |
| 10 | 11 | -10.67 | 85.95 | 42.73 | < .001 |
| 11 | 12 | -12.32 | 84.30 | 44.31 | < .001 |
| 12 | 13 | -6.24 | 90.38 | 38.50 | < .001 |
| **13** | 14 | 7.83 | 104.45 | 25.03 | .09 |

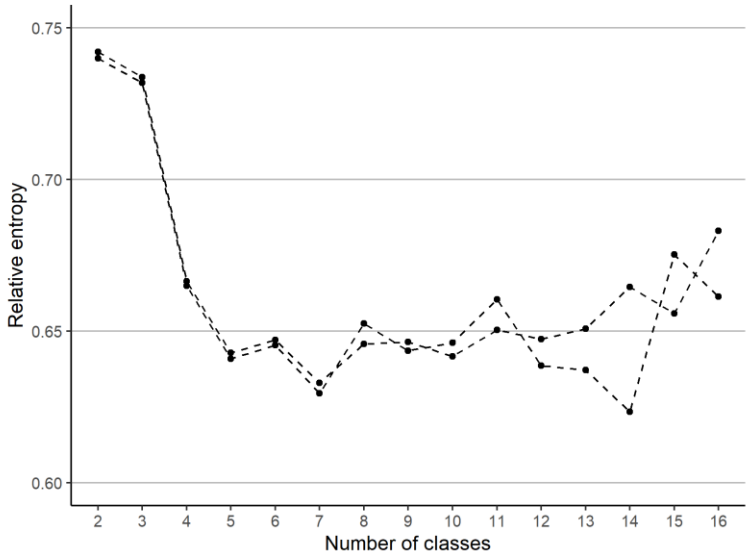


*Figure S2.1. Relative entropy for each estimated model. The entropy for all cases and for all cases except 8 patients with imputed age are shown.*

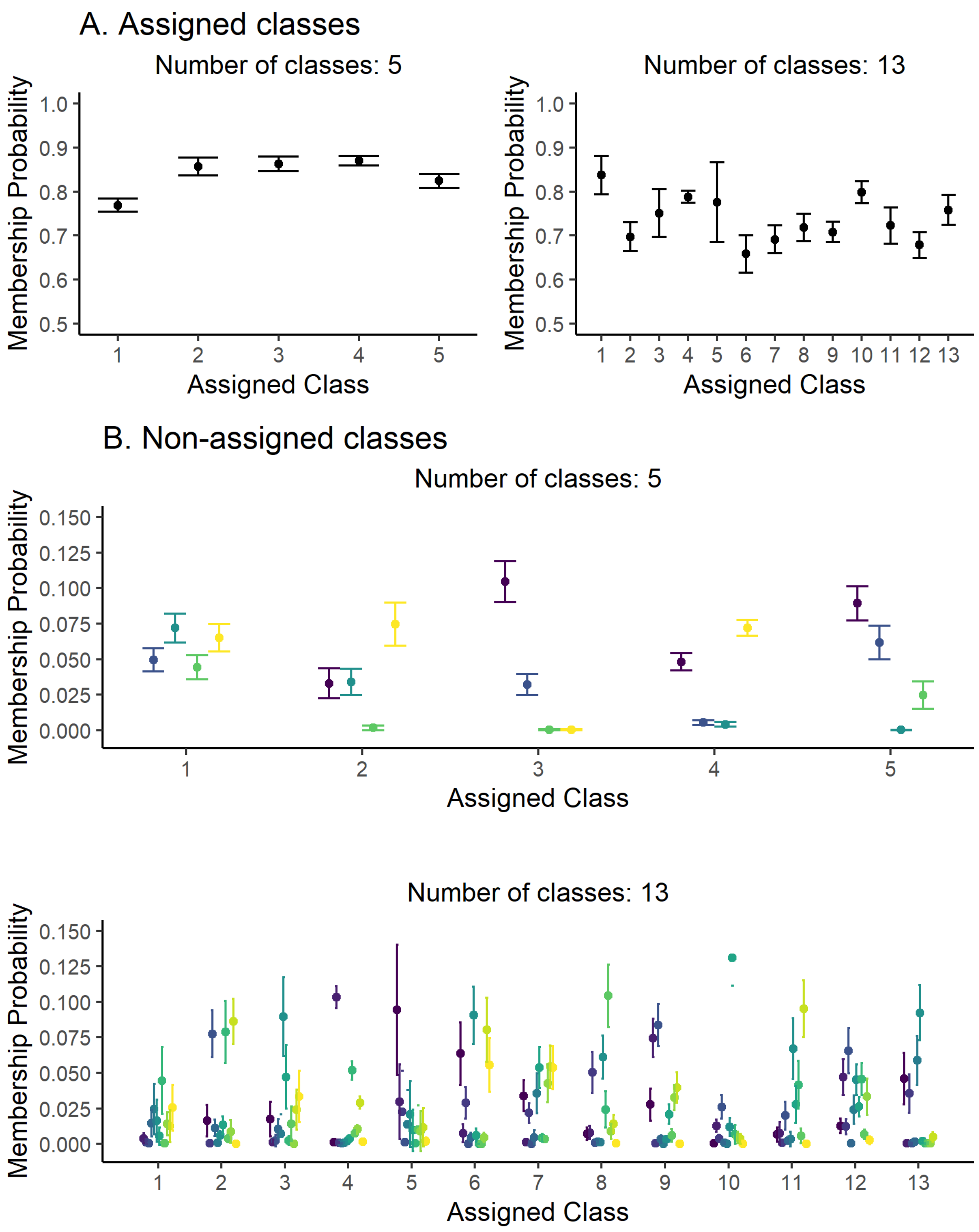


*Figure S2.2. Membership probabilities for each class (separate error bars) for the assigned classes (A) and for the non-assigned classes (B) for the 5-class and 13-class solution.*

#### Additional information 13-class solution

Classes differed in the number of impaired OCS subtests (Table S2.2). The probability of impairment on each OCS subtest for each class is visualized in Figure S2.3. For each subtest the classes are ordered according to the probability of an impaired performance on the subtest. These results reveal that each task contributed to the classification, as none of the tasks are equivalent in impairment probability between the 13 classes.

**Table S2.2. Expected number of impaired subtests for each class**

| Class | M | 95% CI | |
|---|---|---|---|
| 4 | 0.61 | 0.55 | 0.67 |
| 2 | 2.30 | 2.06 | 2.55 |
| 9 | 2.88 | 2.65 | 3.12 |
| 10 | 3.89 | 3.61 | 4.19 |
| 12 | 4.00 | 3.73 | 4.28 |
| 11 | 4.52 | 4.13 | 4.92 |
| 8 | 6.04 | 5.69 | 6.39 |
| 1 | 7.01 | 6.52 | 7.50 |
| 5 | 7.30 | 6.33 | 8.26 |
| 6 | 7.78 | 7.24 | 8.31 |
| 7 | 8.06 | 7.69 | 8.43 |
| 3 | 8.98 | 8.39 | 9.56 |
| 13 | 11.58 | 11.19 | 11.98 |

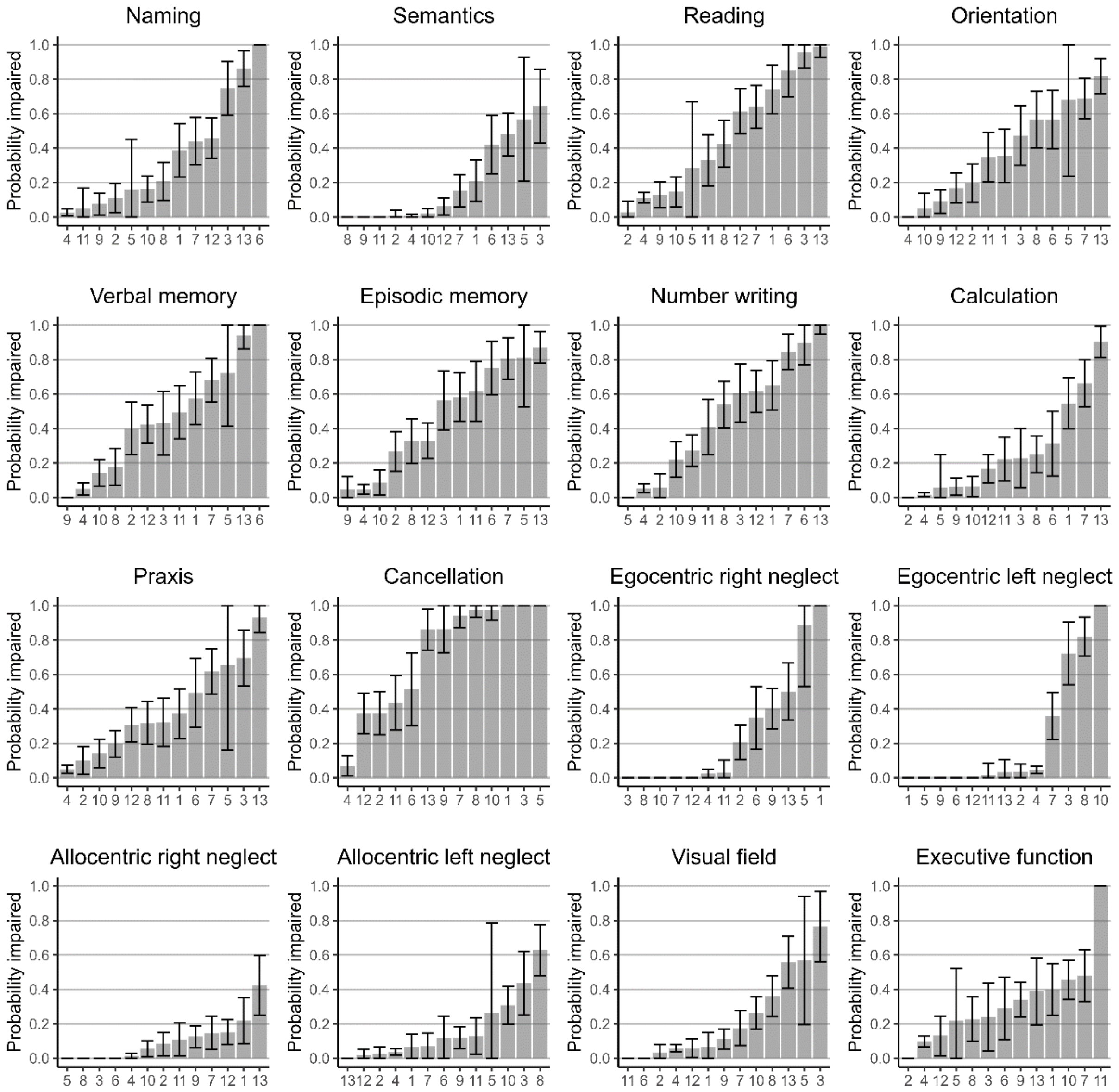


*Figure S2.3. Probability of impairment per class on each OCS subtest sorted in ascending order per panel. The error bar represents the 95% confidence interval.*

### Demographic characteristics of classes

#### Method of analysis

To account for the uncertainty of the class memberships (Bakk & Kuha, 2021), we estimated the probability of observing each demographic covariate level (i.e., age, education, sex, time since stroke and lesion side) given a certain latent class using a Bayesian multinomial logistic regression model that considered the measurement error of the latent class predictor (similar to the approach described in Elliott et al., 2020). The Bayesian measurement error regression model was implemented in Stan (Stan Development Team, n.d.) and fitted with RStan (Stan Development Team, 2020). Then, based on the estimates, the expected difference and their corresponding 99% credible intervals in proportions per covariate level were computed between members of the class versus all others.

For lesion volume a linear regression model was used with lesion volumes log-transformed to correct for their skewness. For the GCA and Fazekas, hierarchical ordinal regression models were used, treating the individual scale items as repeated within-subject measures. The models for lesion volume, GCA and Fazekas ratings were fit using the R package brms (Bürkner, 2017) with default priors, 4 chains and 2000 iterations per chain. All regression models converged with Rhat values smaller than 1.05 and good mixing of traces. The model fit was evaluated through graphical posterior checks with the R BayesPlot package (Gabry & Mahr, 2024).

Since a complete cases analysis is associated with a high risk of bias when missing data is not at random (Buuren & Groothuis-Oudshoorn, 2011), we used multivariate imputation by chained equations to impute missing data using information from all available variables in the dataset (Buuren & Groothuis-Oudshoorn, 2011). Predictors to impute missing covariates were determined based on the proportion of usable cases (min. 20%) and the spearman rho correlation (> .10 or < -.10) (Buuren & Groothuis-Oudshoorn, 2011). The resulting predictor matrix is visualized in Figure S2.4. The predictive mean matching method was used to impute missing observations for all subtest scores, age, years of formal education and days since stroke (Buuren & Groothuis-Oudshoorn, 2011). For all categorical variables, a logistic regression or multinomial logistic regression model was used depending on the number of categories (Buuren & Groothuis-Oudshoorn, 2011). The convergence of the imputation model was visually checked using trace plots. All chains mixed, thus showing good convergence. In addition, distributions of imputed and observed values were visually inspected for impossible values resulting from the imputation.

We created 5 imputed datasets and performed the analysis assessing the relationship between demographic variables and LCA clusters on each imputed dataset and the original complete-cases dataset. The estimates were highly similar between all datasets (Figure S2.5).

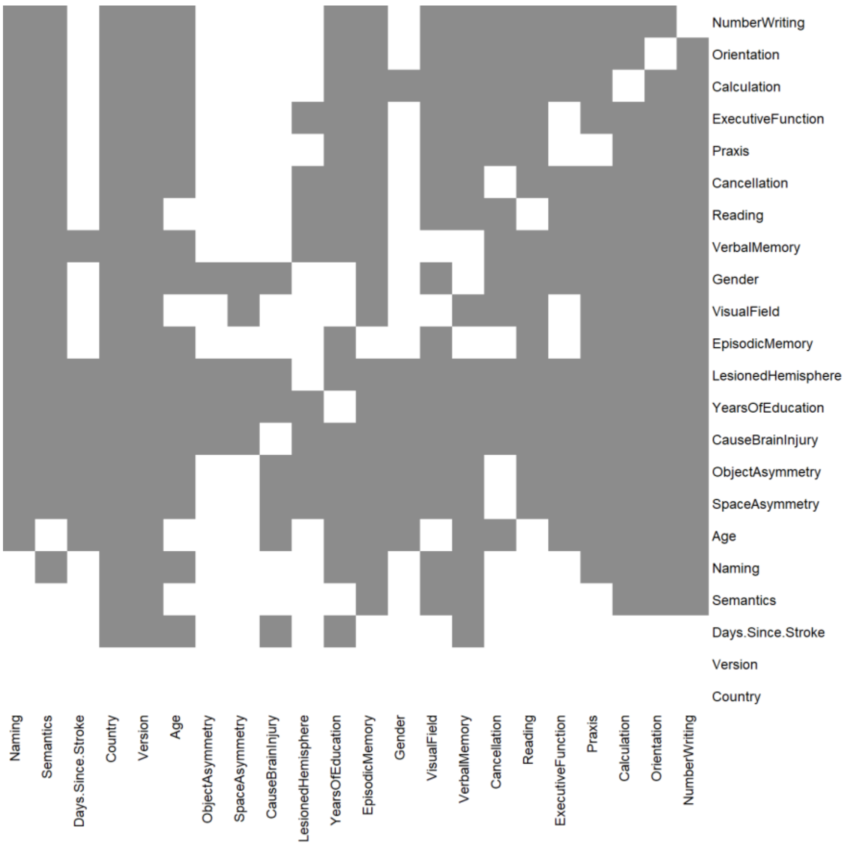


*Figure S2.4. The predictor matrix. A grey square indicates that the variable in the column is used to predict the variable in the row.*

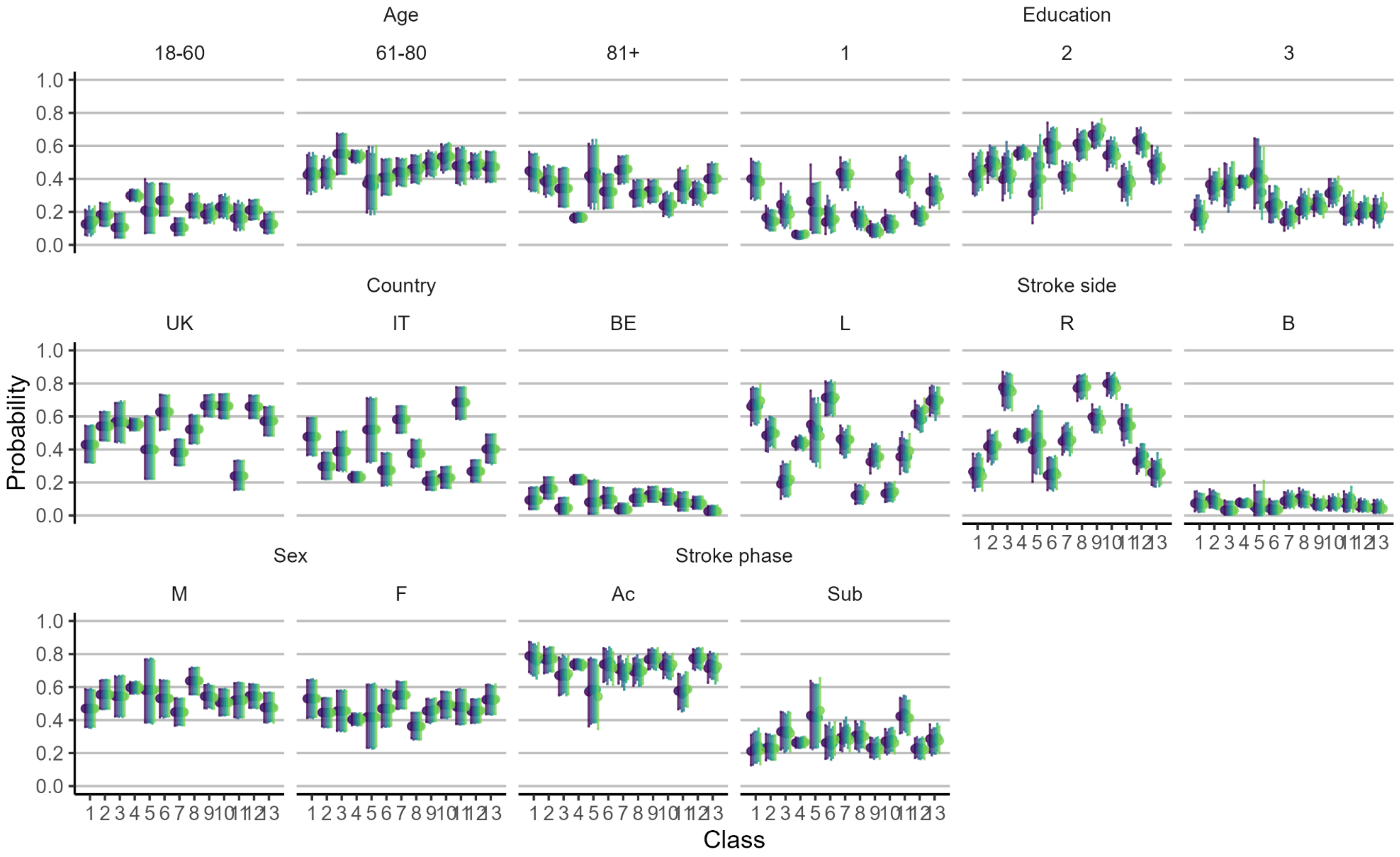


*Figure S2.5. Point estimates and 95% credible intervals for the probability of observing a specific outcome given the latent class for the dataset with complete cases and 5 imputed datasets (different coloured bars).*

***Table S2.3. Participant characteristics for each assigned class.***

| | | ***Age*** | | | | | ***Days since stroke*** | | | | | ***Education*** | | | | | ***Country (%)*** | | | ***Stroke side (%)*** | | | ***Sex (%)*** | |
|---|---|---|---|---|---|---|---|---|---|---|---|---|---|---|---|---|---|---|---|---|---|---|---|---|
| *Class* | *n* | *M* | *Mdn* | *SD* | *Range* | | *M* | *Mdn* | *SD* | *Range* | | *M* | *Mdn* | *SD* | *Range* | | *UK* | *IT* | *BE* | *L* | *R* | *B* | *F* | *M* |
| 1 | 72 | 75.53 | 79.00 | 11.83 | 31 | 92 | 19.26 | 8.00 | 33.96 | 1 | 205 | 8.52 | 9.00 | 3.82 | 3 | 20 | 43 | 49 | 08 | 68 | 26 | 6 | 53 | 47 |
| 2 | 129 | 74.23 | 77.00 | 13.02 | 26 | 92 | 16.11 | 5.00 | 29.98 | 1 | 218 | 11.13 | 11.00 | 4.01 | 3 | 23 | 56 | 29 | 16 | 49 | 42 | 9 | 44 | 56 |
| 3 | 64 | 73.45 | 75.50 | 13.45 | 18 | 95 | 22.19 | 7.50 | 31.55 | 0 | 125 | 10.37 | 11.00 | 4.09 | 3 | 18 | 58 | 39 | 03 | 19 | 79 | 2 | 45 | 55 |
| 4 | 775 | 67.15 | 69.00 | 13.18 | 24 | 96 | 16.47 | 7.00 | 27.06 | 0 | 423 | 12.04 | 12.00 | 3.76 | 3 | 30 | 55 | 23 | 22 | 44 | 48 | 08 | 40 | 60 |
| 5 | 22 | 72.50 | 72.50 | 12.09 | 46 | 90 | 38.16 | 10.00 | 71.25 | 1 | 293 | 10.50 | 9.50 | 4.60 | 5 | 18 | 41 | 55 | 5 | 59 | 41 | 0 | 41 | 59 |
| 6 | 78 | 70.41 | 72.50 | 15.44 | 31 | 94 | 28.24 | 8.00 | 58.85 | 1 | 323 | 10.80 | 10.50 | 3.66 | 5 | 23 | 63 | 28 | 9 | 72 | 25 | 3 | 46 | 54 |
| 7 | 149 | 76.85 | 79.00 | 12.23 | 24 | 97 | 23.90 | 11.00 | 40.28 | 0 | 285 | 8.17 | 8.00 | 4.27 | 1 | 25 | 39 | 58 | 3 | 47 | 44 | 9 | 55 | 45 |
| 8 | 137 | 71.11 | 73.00 | 14.17 | 21 | 93 | 20.71 | 6.00 | 34.53 | 0 | 235 | 10.09 | 10.00 | 3.47 | 3 | 23 | 53 | 37 | 9 | 12 | 78 | 10 | 36 | 64 |
| 9 | 197 | 72.28 | 73.00 | 12.65 | 23 | 93 | 17.74 | 7.00 | 30.93 | 0 | 273 | 10.59 | 10.00 | 3.11 | 4 | 20 | 66 | 22 | 12 | 33 | 60 | 7 | 46 | 54 |
| 10 | 158 | 70.72 | 72.00 | 12.58 | 34 | 96 | 23.79 | 9.00 | 58.41 | 0 | 567 | 10.94 | 11.00 | 3.71 | 3 | 20 | 66 | 23 | 11 | 13 | 81 | 6 | 49 | 51 |
| 11 | 85 | 74.41 | 77.00 | 11.77 | 29 | 93 | 24.44 | 14.00 | 32.22 | 0 | 199 | 8.35 | 8.00 | 3.92 | 3 | 23 | 25 | 69 | 6 | 36 | 58 | 7 | 48 | 52 |
| 12 | 189 | 71.60 | 74.00 | 13.25 | 25 | 94 | 14.12 | 5.00 | 21.89 | 0 | 181 | 9.83 | 10.00 | 3.41 | 2 | 20 | 66 | 28 | 7 | 61 | 34 | 5 | 45 | 55 |
| 13 | 117 | 75.95 | 78.00 | 11.87 | 40 | 98 | 27.80 | 11.00 | 45.51 | 1 | 235 | 8.84 | 9.00 | 3.75 | 2 | 18 | 58 | 40 | 2 | 70 | 26 | 4 | 53 | 47 |

#### Relation between 5- and 13-class solution

We examined how the two class solutions were related to each other (Figure 2.6). Some classes from solution 13 had strong associations with solution 5. That is, 70% of the class 1 members of the 13-class solution were members of class 1 in the 5-class solution. 90% of class 4 members were also members of class 4 in the 5-class solution. 91% of class 8 members were members of class 2 in the 5-class solution. 76% of class 9 were members of class 5 in the 5-class solution. And 88% of class 11 and class 12 were members of class 1 in the 5-class solution. Last, all members of class 13 were members of class 3 in the 5-class solution. Other classes of the 13-class solution had a weaker link to the classes in the 5-class solution, as they consist of mixes of at least 2 classes of the 5-class solution.

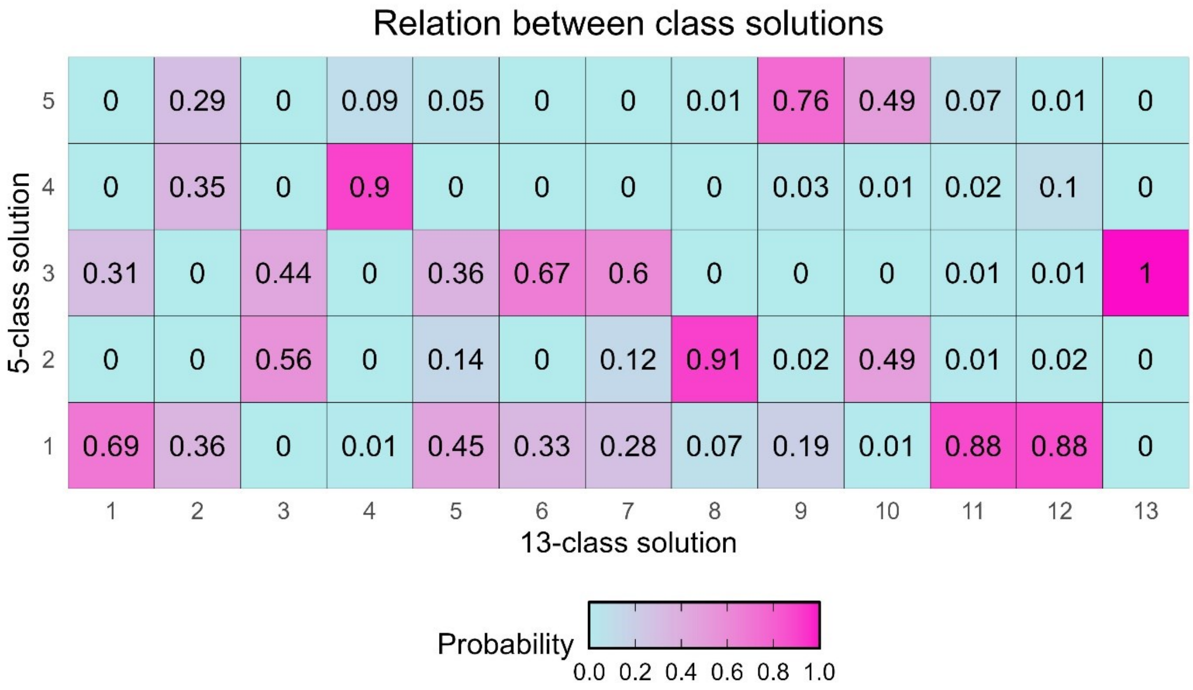


*Figure S2.6. The probability that a patient belongs to a class of the 5-class solution given their membership in the 13-class solution.*

### Supplementary Materials 3. Neural data-analysis

#### Association of cognitive profiles and lesion side

The lesion side was derived from radiologist reports and was available for 1750 (81%) patients. Missing data were imputed according to the procedure described above and several imputed datasets were analysed to assess the impact of imputation (Figure S2.6). The association between behavioural classes and lesion side was examined with the same statistical approach as used for the demographic covariates of class members.

#### Association of cognitive profiles and lesion volume

Patients in Class 3 had the largest average lesion volume (Table S3.1). Lesion volumes varied considerably between patients assigned to a behavioural class (Table S3.1).

**Table S3.1. Lesion volume per class in cubic cm**

| Class | n | M | SD | Med | Min | Max |
|---|---|---|---|---|---|---|
| 1 | 13 | 30.76 | 47.96 | 7.94 | 0.22 | 174.22 |
| 2 | 32 | 20.59 | 24.55 | 13.28 | 0.37 | 114.99 |
| 3 | 17 | 128.83 | 131.10 | 97.49 | 3.06 | 444.46 |
| 4 | 166 | 21.45 | 38.68 | 6.01 | 0.05 | 306.67 |
| 5 | 4 | 25.96 | 33.18 | 11.41 | 5.66 | 75.37 |
| 6 | 25 | 55.73 | 52.62 | 41.67 | 0.06 | 167.69 |
| 7 | 22 | 36.71 | 52.35 | 12.73 | 1.37 | 224.43 |
| 8 | 37 | 57.90 | 60.89 | 38.42 | 0.38 | 231.82 |
| 9 | 50 | 21.12 | 23.43 | 12.52 | 0.18 | 123.34 |
| 10 | 57 | 50.12 | 57.66 | 35.14 | 0.50 | 265.66 |
| 11 | 9 | 7.96 | 8.40 | 2.31 | 0.07 | 21.80 |
| 12 | 51 | 35.03 | 48.90 | 16.68 | 0.02 | 208.35 |
| 13 | 31 | 55.73 | 89.28 | 29.61 | 0.08 | 411.66 |

#### Association of cognitive profiles and lesion location

To assess the association of classes and lesion location, a one-sided Liebermeister test was performed in NiiStat (i.e., testing the hypothesis that a lesion in a voxel increases the probability of belonging to a certain class), controlling for lesion volume as a covariate. Voxels damaged in less than 10 patients were excluded from analyses. Before performing the analysis we checked the representativeness of the patients with lesion maps for the total dataset and we performed a sensitivity analysis.

#### Stability of class solution for patients with and without lesion maps

We first assessed to what extent the cognitive profiles of the 13 behavioural classes were similar in the subset of patients with a lesion map versus the subset of patients without a lesion map (Figure S3.1). For most behavioural classes, the proportion of patients with an impairment are similar between both subsets (Figure S3.1).

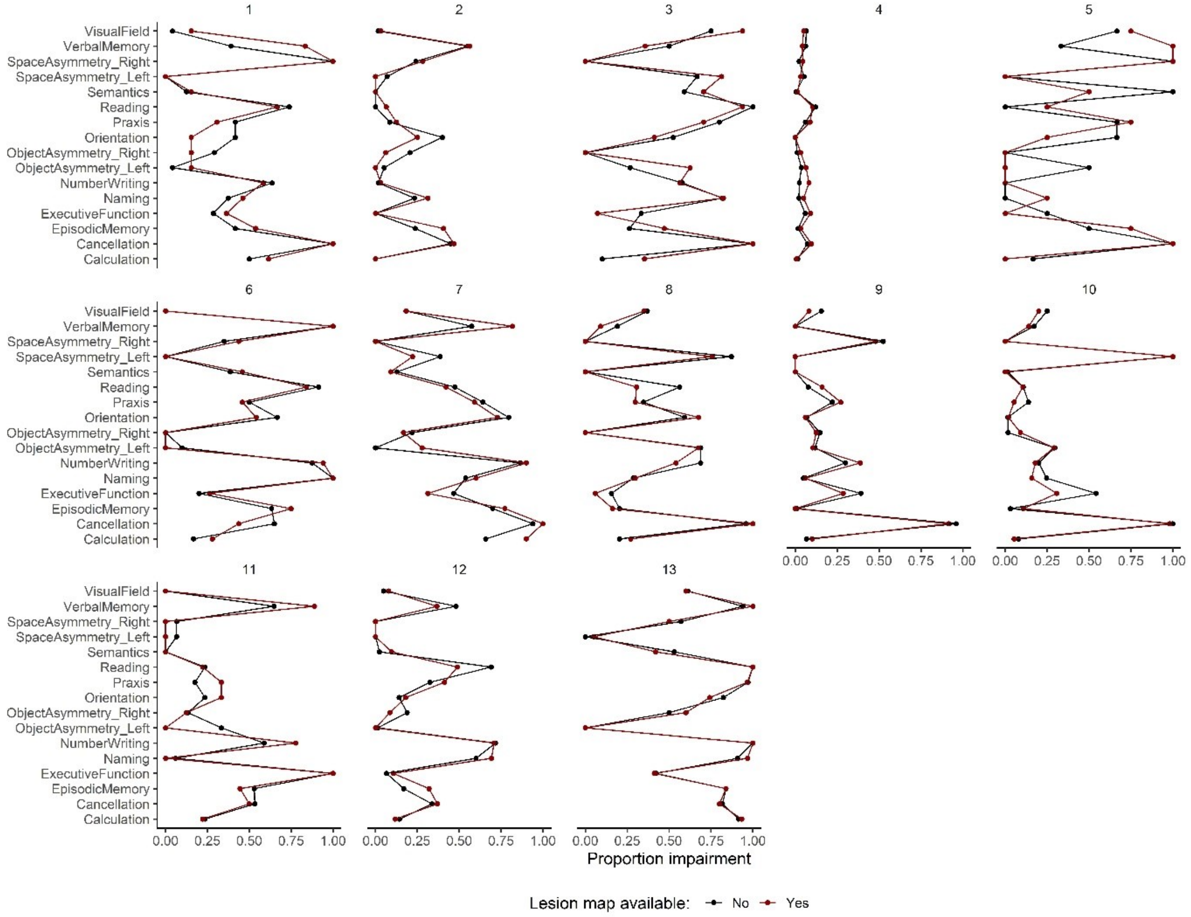


*Figure S3.1. Cognitive profile of behavioural class for patients with versus without a lesion map.*

#### Power and sensitivity analysis

We conducted a sensitivity analysis to determine which effect sizes we could identify with at least 80% power given our sample size. This information provides insight in the sensitivity of our VLSM analysis.

##### *Method*

We first determined the threshold for a voxel to be considered statistically significantly associated to behavioural impairment at a family-wise error rate of 5% using NiiStat. We therefore performed a VLSM including all voxels in the analysis in which at least 5 patients had a lesion and using permutation correction with 2000 permutations with lesion volume as a covariate. This analysis resulted in 141,922 voxels to be included in the VLSM analysis and a critical z-score ranging from -4.49 to -4.72 depending on the behavioural class. We then determined the alpha level corresponding to a statistical test of a single voxel based on the minimum and maximum critical z-scores (alpha: 0.000002 and 0.000004). A single voxel is considered statistically significantly associated with behavioural impairment when the p-value is lower than this alpha level.

We then simulated contingency tables based on four parameters. (1) The total number of patients (N). This parameter was fixed to our sample size (n = 515). (2) The number of patients who have a behavioural impairment (k) (i.e., belong to a specific behavioural class). The simulated range of k was based on the sizes of the behavioural classes for which lesion maps were available and ranged from 5 to 50. (3) The number of patients who have a lesion in a voxel (n). The range of n was based on the maximum lesion overlap in our sample (n = 80) and the minimum lesion overlap for voxels to be included in the VLSM (n = 5). (4) The last parameter was the number of patients who have a lesion in the voxel and a behavioural impairment (q). Q ranged from 1 to a maximum equal to all patients with a behavioural impairment or all patients with a lesion in a voxel. Moreover, only contingency tables where the odds ratio was above or equal to 1 were simulated, as this represents a higher probability of behavioural impairment when a patient has a lesion (i.e., corresponding to the one-sided hypothesis under examination).

A total of 1000 contingency tables were simulated for each set of parameters. For each simulated contingency table, the p-value was calculated. We used the one-sided Liebermeister test to determine the p-value for the relation between behavioural impairment (binary) and lesion in a voxel (binary). The p-value was directly inferred from the hypergeometric distribution using four parameters (i.e., n+1, (N-n)+1, k+1 and q+1). Then, the proportion of significant p-values across the 1000 simulated tables was computed (i.e., power).

##### *Results*

The results reveal that the number of patients with a behavioural impairment and a lesion in the voxel affect the sensitivity of the VLSM analysis (Table S3.2, Figure S3.2). The highest sensitivity (lowest detectable odds ratio) is achieved when both or equal to 50 (Table S3.2). Moreover, the simulation revealed that for rare behavioural impairments (k=5), we only reach 80% power if 80% to 100% of the patients with the behavioural impairment also show a lesion in the voxel (Figure S3.3). The latter corresponds to very large odds ratios (OR>1000, Table S3.2). Similarly, for voxels that are only lesioned in 5 patients, 80% power could not always be obtained (Figure S3.2, Table S3.2).

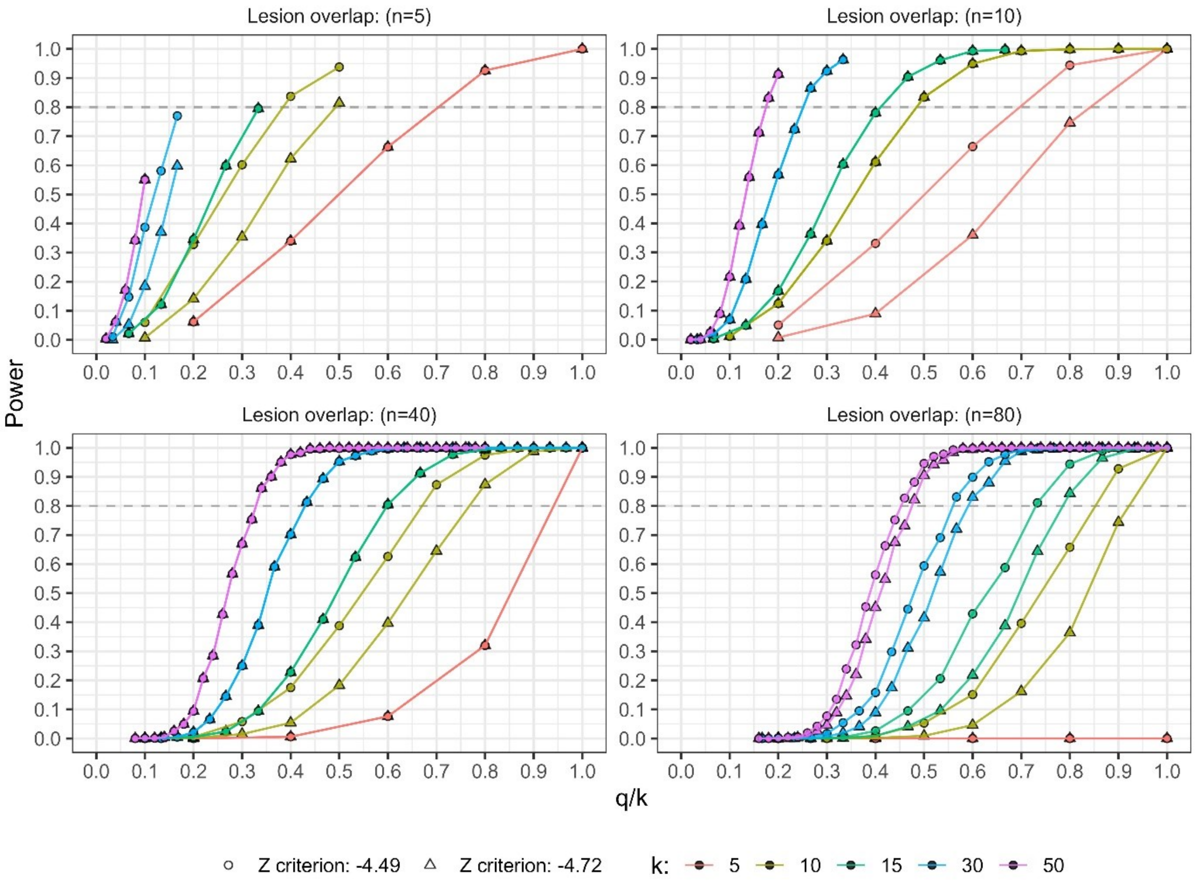


*Figure S3.2. Power curves for VLSM. The achieved power for different contingency tables characterized by a number of patients with behavioural impairment (k), number of patients with lesion overlap in the voxel (n) and proportion of patients with behavioural impairment and a lesion relative to number of patients with behavioural impairment (q/k) for the two critical z-scores.*

**Table S3.2. Odds ratios and (achieved power) for different sample sizes of behavioural impairment and lesion overlap.**

| | | Lesion overlap (n) | | | | | | |
|---|---|---|---|---|---|---|---|---|
| | | **5** | **10** | **20** | **30** | **40** | **50** | **80** |
| Behavioural impairment (n) | **5** | >1000 (.95) | >1000 (1.00) | >1000 (1.00) | >1000 (1.00) | >1000 (1.00) | >1000 (1.00) | * |
| | **10** | >1000 (.85) | 100 (.82) | 53 (.83) | 49 (.84) | 59 (.88) | 44 (.88) | >1000 (1.00) |
| | **15** | * | 145 (.91) | 46 (.88) | 34 (.90) | 31 (.92) | 23 (.81) | 25 (.84) |
| | **30** | * | 88 (.85) | 24 (.82) | 17 (.82) | 13 (.84) | 11 (.82) | 10 (.83) |
| | **50** | * | 102 (.82) | 18 (.80) | 13 (.86) | 10 (.86) | 9 (.83) | 7 (.82) |

Table Note. * = 80% power not achieved.
All estimates for power and odds ratios are for a critical z-score = -4.72.

##### *Conclusion*

Given the sensitivity analysis, we only performed the VLSM for the behavioural classes for which at least 15 patients had a lesion map and for which there was overlap of lesions in at least 10 patients. The sensitivity analysis revealed that the minimal odds ratio that we can detect with more than 80% power given these criteria equals 7. Small odds ratios can thus not be detected.

#### Results of VLSM and atlas comparison

Resultant significant voxels clusters (>10 contiguous voxels) were compared to anatomical atlases using the Python package AtlasReader (Notter et al., 2019). Neuroanatomical data were visualized using nilearn (Abraham et al., 2014).

##### *5-class solution*

Table S3.3. Clusters of significant voxels, their mean intensity, volume and overlap with anatomical regions from three atlases for the 5-class solution.

| | MI | Vol ($mm^3$) | AAL atlas | Desikan-Kiliany atlas | Harvard Oxford atlas |
|---|---|---|---|---|---|
| **Class 2** | | | | | |
| 1 | -5 | 150248 | 23.35% no label<br>11.26% Temporal Sup R<br>9.05% Temporal Mid R<br>7.11% Insula R<br>6.99% Angular R<br>6.08% Rolandic Oper R | 46.31% R CWM<br>17.59% Unknown<br>5.64% R inferior parietal | 10.86% R LOC superior division<br>7.97% R Insular Cortex<br>6.28% R Angular Gyrus<br>5.92% R Central Opercular Cortex<br>5.62% R Middle Temporal Gyrus temporo-occipital part |
| 2 | -4.54 | 128 | 62.50% Frontal Inf Oper R<br>18.75% Frontal Inf Tri R<br>18.75% no label | 93.75% R CWM<br>6.25% R rostral middle frontal | 100.00% R Middle Frontal Gyrus |
| **Class 3** | | | | | |
| 1 | -4.94 | 16280 | 36.17% Insula L<br>31.25% no label<br>11.45% Putamen L<br>7.62% Frontal Inf Oper L | 45.11% L CWM<br>17.25% L Insula<br>17.25% Unknown<br>12.73% L Putamen | 38.48% L Insular Cortex<br>17.69% L Putamen<br>11.15% L Central Opercular Cortex<br>7.62% no label<br>7.57% L Frontal Operculum Cortex<br>7.37% L Precentral Gyrus |
| 2 | -4.63 | 352 | 100.00% Precentral L | 45.45% Unknown<br>36.36% L precentral<br>18.18% L CWM | 68.18% L Precentral Gyrus<br>31.82% L Middle Frontal Gyrus |
| 3 | -4.67 | 240 | 100.00% Frontal Inf Tri L | 40.00% Unknown<br>36.67% L rostral middle frontal<br>23.33% L CWM | 100.00% L Middle Frontal Gyrus |
| 4 | -4.79 | 112 | 100.00% no label | 100.00% L CWM | 42.86% no label<br>35.71% L Precuneus<br>21.43% L Lateral Ventrical |

Table S3.4. Coordinates and peak intensity of peak values and their corresponding anatomical labels for the 5-class solution.

| class | cluster | x | y | z | PI | AAL | Desikan-Kiliany | Harvard Oxford |
|---|---|---|---|---|---|---|---|---|
| 2 | 1 | 46 | -4 | 2 | -7.70 | Insula R | Unknown | 21.0% R Central Opercular Cortex<br>20.0% R Insular Cortex<br>8.0% R Heschl's Gyrus<br>6.0% R Planum Polare |
| | 2 | 30 | 14 | 30 | -4.83 | Frontal Inf Oper R | R CWM | 10.0% R Middle Frontal Gyrus |
| 3 | 1 | -32 | 4 | 8 | -6.69 | Insula L | L CWM | 39.0% L Insular Cortex |
| | 2 | -46 | 0 | 42 | -5.08 | Precentral L | L CWM | 49.0% L Precentral Gyrus<br>10.0% L Middle Frontal Gyrus |
| | 3 | -40 | 24 | 26 | -5.21 | Frontal Inf Tri L | L rostral middle frontal | 39.0% L Middle Frontal Gyrus<br>9.0% L Inferior Frontal Gyrus pars triangularis |
| | 4 | -28 | -60 | 18 | -5.08 | no label | L CWM | 0% no label |

#### *13-class solution*

***Table S3.5. Clusters of significant voxels per class, their mean intensity (average z-score), volume and their overlap with anatomical regions from three atlases for the 13-class solution.***

| | *MI* | *Vol ($mm^3$)* | *AAL atlas* | *Desikan-Kiliany atlas* | *Harvard Oxford atlas* |
|---|---|---|---|---|---|
| | | | | **Class 3** | |
| 1 | -4.94 | 70,144 | 24.68% no label<br>15.07% Calcarine R<br>14.30% Temporal Mid R<br>10.52% Lingual R<br>7.66% Temporal Inf R<br>7.44% Temporal Sup R | 58.90% R CWM<br>10.66% Unknown | 11.25% R LOC inferior division<br>10.62% R Middle Temporal Gyrus posterior division<br>10.13% R Intracalcarine Cortex<br>8.03% R Lingual Gyrus<br>7.04% R Middle Temporal Gyrus temporooccipital part<br>6.50% R Occipital Fusiform Gyrus<br>5.54% R Occipital Pole |
| 2 | -4.42 | 864 | 55.56% no label<br>27.78% Frontal Inf Oper R<br>15.74% Frontal Mid 2 R | 58.33% R CWM<br>22.22% R caudal middle frontal<br>16.67% Unknown | 37.04% no label<br>31.48% R Middle Frontal Gyrus<br>24.07% R Precentral Gyrus<br>6.48% R Inferior Frontal Gyrus pars opercularis |
| 3 | -4.49 | 320 | 75.00% no label<br>12.50% Amygdala R<br>10.00% Hippocampus R | 40.00% R Amygdala<br>30.00% R Ventral DC<br>17.50% R CWM<br>10.00% R Pallidum | 57.50% R Amygdala<br>40.00% R Pallidum |
| 4 | -4.31 | 288 | 88.89% Precentral R<br>11.11% Frontal Inf Oper R | 47.22% R CWM<br>41.67% R precentral<br>11.11% Unknown | 94.44% R Precentral Gyrus<br>5.56% R Inferior Frontal Gyrus pars opercularis |
| 5 | -4.37 | 288 | 94.44% Supramarginal R<br>5.56% Temporal Sup R | 72.22% R supramarginal<br>13.89% Unknown<br>11.11% R CWM | 50.00% R Supramarginal Gyrus anterior division<br>44.44% R Supramarginal Gyrus posterior division<br>5.56% R Parietal Operculum Cortex |

| | | | | | |
|---|---|---|---|---|---|
| 6 | -4.37 | 272 | 73.53% Frontal Inf Oper R<br>17.65% Rolandic Oper R 8.82% Frontal Inf Tri R | 52.94% Unknown<br>29.41% R pars opercularis<br>14.71% R precentral | 79.41% R Inferior Frontal Gyrus pars opercularis<br>20.59% R Precentral Gyrus |
| 7 | -4.49 | 152 | 84.21% no label<br>10.53% Putamen R<br>5.26% Insula R | 100.00% R CWM | 52.63% R Insular Cortex<br>36.84% no label<br>10.53% R Putamen |
| 8 | -4.44 | 144 | 100% Hippocampus R | 100% R Hippocampus | 100 % R Hippocampus |
| 9 | -4.36 | 128 | 100% Putamen R | 100% R Putamen | 100 % R Putamen |
| | | | **Class 6** | | |
| 1 | -5.13 | 28832 | 31.35% Insula L<br>30.77% no label<br>12.35% Frontal Inf Oper L<br>10.16% Putamen L<br>5.16% Frontal Inf Tri L | 38.40% L CWM<br>22.23% Unknown<br>14.73% L insula<br>11.65% L Putamen<br>6.44% L pars opercularis | 32.55% L Insular Cortex<br>15.90% L Putamen<br>10.38% L Inferior Frontal Gyrus pars opercularis<br>9.54% L Frontal Operculum Cortex<br>8.66% no label<br>7.33% L Central Opercular Cortex<br>5.44% L Precentral Gyrus |
| 2 | -4.71 | 304 | 100.00% no label | 100.00% L CWM | 78.95% no label<br>15.79% L Insular Cortex<br>5.26% L Central Opercular Cortex |
| | | | **Class 8** | | |
| 1 | -4.70 | 624 | 61.54% Insula R<br>38.46% Temporal Sup R | 67.95% Unknown<br>16.67% R insula<br>11.54% R superior temporal | 46.15% R Insular Cortex<br>29.49% R Heschl's Gyrus<br>23.08% R Planum Polare |
| | | | **Class 10** | | |
| 1 | -4.83 | 3880 | 63.30% no label<br>16.70% Thalamus R<br>15.46% Putamen R | 68.45% R CWM<br>16.29% R Thalamus Proper<br>9.48% R Putamen | 36.08% R Putamen<br>31.55% R Thalamus<br>20.00% no label |
| 2 | -4.64 | 80 | 60.00% Postcentral R<br>40.00% no label | 40.00% ctx R precentral<br>40.00% R CWM<br>20.00% Unknown | 70.00% R Precentral Gyrus<br>30.00% R Postcentral Gyrus |
| 3 | -4.53 | 80 | 100% no label | 90.00% R CWM<br>10.00% R precentral | 100% R Postcentral Gyrus |
| | | | **Class 12** | | |
| 1 | -4.48 | 192 | 75.00% Heschl L<br>16.67% Temporal Sup L<br>8.33% Insula L | 50.00% Unknown<br>20.83% L insula<br>16.67% L transverse temporal<br>12.50% L CWM | 66.67% L Heschl's Gyrus<br>33.33% L Insular Cortex |

| | | | | | |
|---|---|---|---|---|---|
| 2 | -4.57 | 176 | 81.82% Temporal Sup L<br>18.18% Insula L | 50.00% Unknown<br>31.82% L superior temporal<br>13.64% L CWM | 86.36% L Planum Polare<br>13.64% L Insular Cortex |
| | | | **Class 13** | | |
| 1 | -4.54 | 552 | 44.93% no label<br>28.99% Precentral L<br>21.74% Frontal Inf Oper L | 71.01% L CWM<br>14.49% L caudal middle frontal<br>10.14% Unknown | 73.91% L Precentral Gyrus<br>15.94% L Middle Frontal Gyrus<br>5.80% no label |
| 2 | -4.56 | 336 | 92.86% Frontal Inf Tri L<br>7.14% no label | 54.76% Unknown<br>40.48% L pars triangularis | 100 % L Inferior Frontal Gyrus pars triangularis |
| 3 | -4.56 | 144 | 55.56% Insula L<br>44.44% no label | 77.78% L insula<br>22.22% L CWM | 100 % L Insular Cortex |
| 4 | -4.54 | 120 | 86.67% Hippocampus L<br>13.33% no label | 46.67% L choroid plexus<br>46.67% L CWM<br>6.67% L Hippocampus | 100 % L Hippocampus |
| 5 | -4.48 | 104 | 100.00% Frontal Inf Tri L | 76.92% Unknown<br>23.08% L pars opercularis | 84.62% L Inferior Frontal Gyrus pars opercularis<br>7.69% L Inferior Frontal Gyrus pars triangularis<br>7.69% L Middle Frontal Gyrus |
| 6 | -4.53 | 96 | 100.00% no label | 66.67% L CWM<br>25.00% L bankssts<br>8.33% Unknown | 58.33% L Middle Temporal Gyrus posterior division<br>25.00% no label<br>16.67% L Superior Temporal Gyrus posterior division |
| 7 | -4.56 | 88 | 100.00% no label | 100% L CWM | 90.91% no label<br>9.09% L Hippocampus |

Note. MI = cluster mean intensity, Vol = lesion volume, LOC = Lateral Occipital Cortex, CWM = Cerebral White Matter.

***Table S3.6. Coordinates and peak intensity of peak values and their corresponding anatomical labels** **for the 13-class solution**.*

| *class* | *cluster* | *x* | *y* | *z* | *PI* | *Atlas comparison* | | |
|---|---|---|---|---|---|---|---|---|
| | | | | | | ***AAL*** | ***Desikan-Kiliany*** | ***Harvard Oxford*** |
| 3 | 1 | 24 | -80 | 6 | -6.92 | / | R CWM | / |
| | 2 | 32 | -6 | 26 | -4.86 | / | R CWM | / |
| | 3 | 20 | -6 | -10 | -5.06 | / | R Amygdala | 44% R Amygdala |
| | 4 | 56 | 4 | 32 | -4.43 | Precentral R | R CWM | 43% R Precentral Gyrus |
| | 5 | 66 | -36 | 30 | -4.34 | SupraMarginal R | R supramarginal | 45% R Supramarginal Gyrus posterior division |
| | 6 | 60 | 16 | 6 | -4.63 | Frontal Inf Oper R | / | 31% R Inferior Frontal Gyrus pars opercularis |
| | 7 | 32 | -2 | 14 | -4.68 | / | R CWM | 7% R Insular Cortex |
| | 8 | 30 | -18 | -16 | -4.99 | Hippocampus R | R Hippocampus | 99% R Hippocampus |
| | 9 | 26 | -4 | 10 | -4.55 | Putamen R | R Putamen | 98% R Putamen |
| 6 | 1 | -30 | 6 | 12 | -7.45 | Insula L | L CWM | 23% L Insular Cortex |
| | 2 | -30 | -20 | 26 | -5.32 | / | L CWM | / |
| 8 | 1 | 48 | -10 | 0 | -5.18 | Temporal Sup R | / | 31% R Heschl's Gyrus<br>24% R Planum Polare |
| 10 | 1 | 22 | -18 | 18 | -5.82 | / | R CWM | / |

| | | | | | | | | |
|---|---|---|---|---|---|---|---|---|
| | 2 | 44 | -10 | 32 | -5.15 | Postcentral R | / | 40% R Precentral Gyrus |
| | 3 | 44 | -16 | 32 | -4.71 | / | R CWM | 21% R Postcentral Gyrus |
| 12 | 1 | -42 | -20 | 8 | -4.85 | Heschl L | L transverse temporal | 67% L Heschl's Gyrus |
| | 2 | -44 | -10 | -6 | -5.09 | Temporal Sup L | / | 58% L Planum Polare |
| 13 | 1 | -34 | 2 | 22 | -4.96 | / | L CWM | / |
| | 2 | -58 | 30 | 10 | -4.89 | / | / | 14% L Inferior Frontal Gyrus pars triangularis |
| | 3 | -36 | 6 | -4 | -4.96 | / | Ctx L insula | 54% L Insular Cortex |
| | 4 | -36 | -22 | -12 | -4.81 | Hippocampus L | L CWM | 29% L Hippocampus |
| | 5 | -58 | 20 | 24 | -4.35 | Frontal Inf Tri L | / | 18% L Inferior Frontal Gyrus pars opercularis |
| | 6 | -40 | -40 | 2 | -4.72 | / | L CWM | / |
| | 7 | -40 | -36 | -6 | -4.44 | / | L CWM | / |

*Table Note. MNI coordinates belonging to a certain ROI per atlas (Harvard-Oxford: highest probable ROI).*
*PI = peak intensity. / = MNI coordinate not labelled in atlas.*

#### Association of cognitive profiles and functional dysconnectivity

##### Method

To compare the functional dysconnectivity networks of patients assigned to a class versus all other patients, we first estimated the networks for each patient using the Matlab Lesion Quantification Toolkit (Griffis et al., 2021). This results in a disconnection matrix containing 135 parcels (regions of interest) and 9045 unique edges (i.e., a disconnection between a pair of two parcels). *This matrix was reduced to only contain the edges that were disconnected in at least 1 patient (1375 edges).*

Then, the linear ANOVA model in the R package NBR (Gracia-Tabuenca & Alcauter, 2020) was used to compare the disconnection matrices of class members to other participants. We tested for differences between class members and others for each edge, assuming that edges that differ between groups are clustered into components (i.e., a subnetwork of connected edges). The latter allows for more statistical power than mass-univariate statistical comparisons (Kim et al., 2014; Zalesky et al., 2010).

To identify suprathreshold edges, a p-value threshold of .01 was used. There are no clear guidelines on which threshold to use to identify suprathreshold edges (Kim et al., 2014; Zalesky et al., 2010). For this reason, we checked the differences in edge weights for edges that were part versus not part of the significant component of edges to check how conservative our analysis was (Figure S3.3). Significant components of edges were identified with the family-wise error p-value for the sum of edge weights of the component, estimated with 1000 permutations.

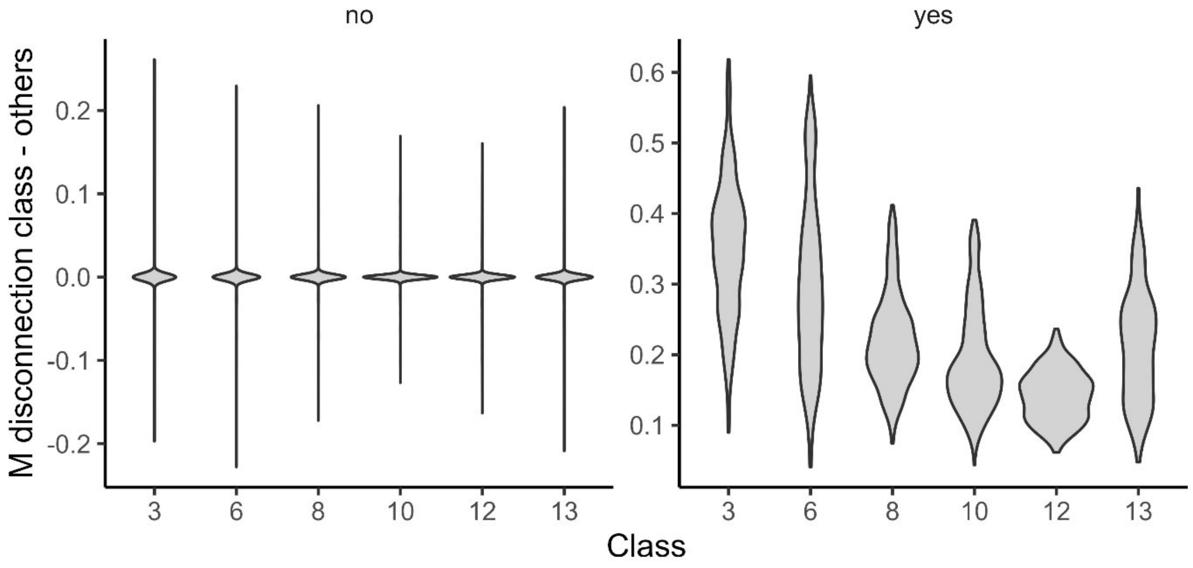


*Figure S3.3. Differences in average disconnections between class members and others for edges not included in the significant network (left panel) and edges included in the significant network (right panel) for each behavioural class for which a significant subnetwork emerged.*

#### Results

For the 5-class solution, a statistically significant subnetwork of disconnections was identified for Class 1, 2, and 3 which consisted of disconnections that were more disconnected in Class members versus all others (Table S3.7). For Class 4, significant subnetwork was identified, but this subnetwork only consisted of disconnections which were less severe in Class 4 members than other patients. Last, for Class 5 not a single significant subnetwork was identified.

For the 13-class solution, a statistically significant subnetwork of disconnections was identified for the right-lateralized classes (i.e., Class 3, 8, and 10), but not for Class 9. For the left-lateralized classes a significant subnetwork was identified for Classes 6, 12 and 13. For the non-lateralized classes, no significant subnetwork consisting of connections that were more disconnected in class members versus others was identified (i.e., Class 2, 4, and Class 7). The other classes (Class 1, 5, and 11) were excluded from the network analysis due to an insufficient number of patients with a lesion map (< 15 patients).

**Table S3.7. Results of the two-sided NBR test**

| Class | Component | FWE corrected p-value edge weights | Number of significant disconnections | |
|---|---|---|---|---|
| | | | Total | Class > Others |
| | | 5 Class Solution | | |
| 1 | 1 | < .001 | 534 | 233 |
| 2 | 1 | < .001 | 1006 | 773 |
| 3 | 1 | < .001 | 550 | 429 |
| 4 | 1 | < .001 | 425 | 0 |

| | | | | |
|---|---|---|---|---|
| | 2 | .55 | 0 | 0 |
| | 3 | .75 | 0 | 0 |
| 5 | 1 | .003 | 156 | 0 |
| | 2 | .29 | 0 | 0 |
| | 3 | .03 | 0 | 0 |
| | 4 | .60 | 0 | 0 |
| | 5 | .75 | 0 | 0 |
| **13 Class Solution** | | | | |
| 2 | 1 | .29 | 0 | 0 |
| 3 | 1 | < .001 | 615 | 360 |
| 4 | 1 | < .001 | 569 | 0 |
| | 2 | .77 | 0 | 0 |
| 6 | 1 | < .001 | 419 | 360 |
| 7 | 1 | .135 | 0 | 0 |
| 8 | 1 | < .001 | 505 | 476 |
| 9 | 1 | .41 | 0 | 0 |
| | 2 | .28 | 0 | 0 |
| | 3 | .79 | 0 | 0 |
| | 4 | .70 | 0 | 0 |
| | 5 | .41 | 0 | 0 |
| 10 | 1 | < .001 | 612 | 493 |
| 12 | 1 | < .001 | 292 | 186 |
| 13 | 1 | < .001 | 394 | 266 |

To gain insight in which functional disconnections were most characteristic of a class, we further inspected the edges which were more disconnected in Class members versus others. We quantified network disconnection as the number of edges which were significantly more disconnected relative to all network edges. An edge was considered part of a functional network if at least one node belonged to the network. Thus, disconnections between two nodes of a functional network and disconnections between a node of a functional network with nodes of other networks are taken into consideration. This approach provides insight in the relative contributions of each functional network for Class membership.

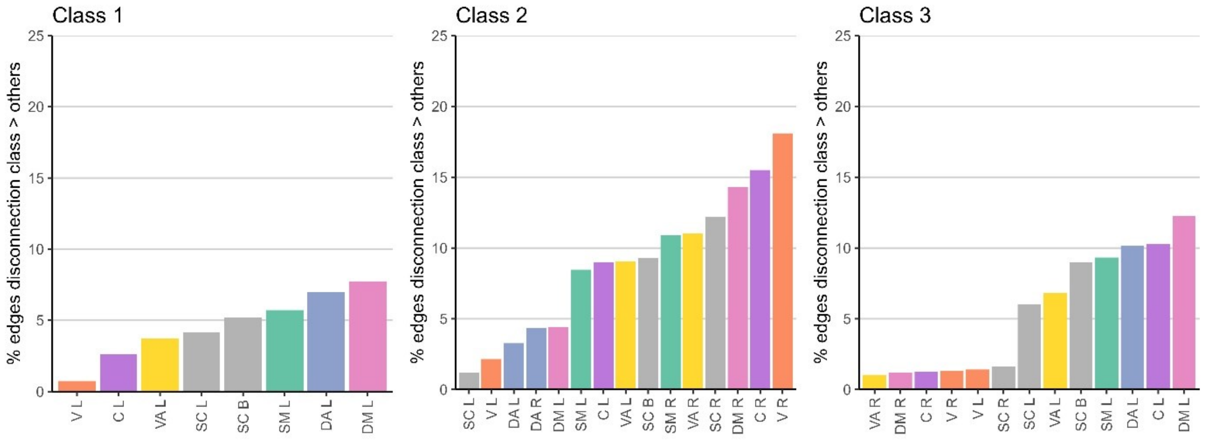


*Figure S3.4. Relative network contributions per class of the 5-class solution for which significant disconnections were identified.*

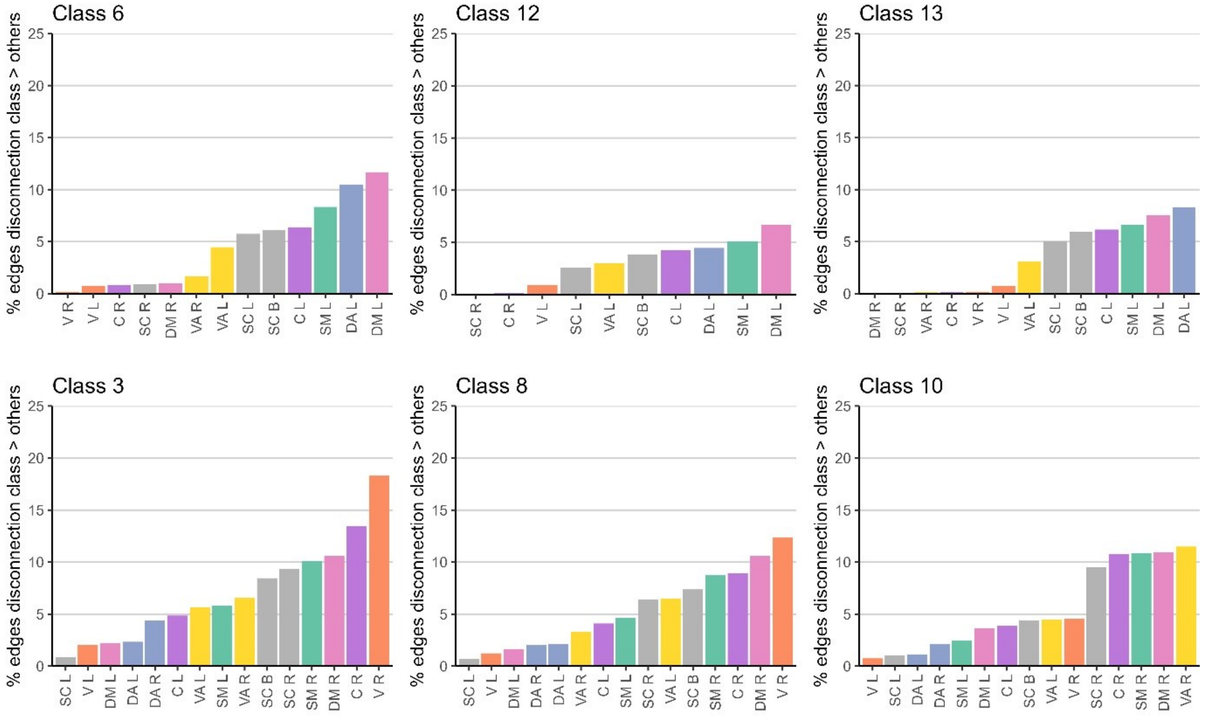


*Figure S3.5. Relative network contributions per class of the 13-class solution for which significant disconnections were identified.*

### Multivariate Similarity Analysis

#### Method

To quantify behavioural similarity, we computed the Manhattan distances between each pair of participants on the class-probability vectors (which are normalized to sum to 1 per participant) extracted from the LCA model.

To quantify neuroanatomical similarity, several metrics were used. Similarity in lesion location at the voxel-level (i.e., lesion overlap) was quantified using the Dice coefficient for each pair of participants. The difference between lesion volume for each pair of participants was also computed.

In addition, neuro-anatomical similarity at the level of disconnections (tract-based and functional disconnections) was quantified. The disconnections were first standardized. Standardizing ensures that features (disconnected tracts or edges) have equal relative weights when computing the distances between each pair of participants. Then, the Manhattan distance metric was calculated for each pair of participants for both matrices separately. The distance metric represents how dissimilar participants are in their disconnections.

We then assessed the pairwise Spearman associations of the behavioural distances with each neuro-anatomical distance (lesion location, lesion volume, and tract and functional disconnections) using the Mantel Test in the R package ecodist (Goslee & Urban, 2007), which considers the non-independence of the distances.

The Mantel Test was estimated for the full sample and for subsets of patients to assess the potential moderating effect of time after stroke, stroke type (ischemic versus haemorrhagic stroke) and severity of premorbid brain health (Fazekas and GCA ratings).

In addition, to assess the potential impact of influential cases, we estimated the associations across subsets of data leaving out one case at a time. This check did not reveal influential observations.
